# Complex intra-host SARS-CoV-2 evolution following monoclonal antibody pre-exposure prophylaxis

**DOI:** 10.64898/2026.07.14.26356329

**Authors:** Kimia Kamelian, David J Pascall, Mark Tsz Kin Cheng, Bo Meng, Mazharul Altaf, Rebecca B Morse, Juliana B Aggio, Dan Egan, Michael Chen-Xu, Giorgio Trivioli, Ben Sutton, Alex Richter, Luis Daniel González Vázquez, Claire Gallagher, Steven A Kemp, Rory Yeadon, Ben Hyatt, Andrew Wong, Nashma Thesin Pelamkulangara, Emma Fraser, Ben McCarthy, Fernanda Novaes, Sara Stott, Anastasia Galvin, Katherine L Bellis, Daniela De Angelis, Ewan M Harrison, Darren Martin, Rona M Smith, Ravindra K Gupta

**Affiliations:** University of Cambridge, School of Clinical Medicine, Department of Medicine, Cambridge, Cambridgeshire, United Kingdom; Cambridge Institute of Therapeutic Immunology & Infectious Disease (CITIID), Cambridge, Cambridgeshire, United Kingdom; MRC Biostatistics Unit, University of Cambridge, East Forvie Building, Cambridge, Cambridgeshire, United Kingdom; Cambridge University Hospitals NHS Foundation Trust, Hills Road, Cambridge, Cambridgeshire, United Kingdom; University Hospitals Birmingham NHS Foundation Trust, Birmingham, United Kingdom; Department of Immunology and Immunotherapy, University of Birmingham, Birmingham, United Kingdom; Biomedical Research Center (CINBIO), University of Vigo, Vigo, Spain; Department of Biochemistry, Genetics and Immunology, University of Vigo, Vigo, Spain; Wellcome Sanger Institute, Wellcome Genome Campus, Hinxton, United Kingdom; Computational Biology Division, Department of Integrative Biomedical Sciences, University of Cape Town, South Africa; Africa Health Research Institute, Durban, South Africa

## Abstract

**Background:** Monoclonal antibodies have emerged as a prophylactic strategy to prevent symptomatic SARS-CoV-2 infection in immunocompromised individuals. However, the evolutionary and clinical implications of breakthrough infections under this regime remain unclear.

**Methods:** A male in their 80s with a haematological/oncological diagnosis received a 2000 mg intravenous infusion of sotrovimab in March 2023 and was diagnosed with COVID-19 by RT-qPCR from a nasopharyngeal swab in August 2023. Weekly samples (n=24) were collected through February 2024 (171 days). All samples underwent whole-genome sequencing, with select mutations subjected to functional assessment.

**Findings:** Sequencing identified the GE.1 lineage at all timepoints. An intra-host recombination event in ORF1ab (positions 8942-12458) was detected prior to 23 weeks post-detection, followed by a 14-fold increase in viral load (7.42e+06 to 1.00e+08 RNA copies/mL) and a marked shift in the viral population. E340D, a sotrovimab resistance mutation, was detected at low abundance (46%) within the first week post-infection, fluctuated over time, and was nearly fixed by week 15 (107 days) post-detection. We assessed five spike mutations – V36M, S98F, and V213G in the N-terminal domain, Y505P in the receptor-binding domain, and P681Q near the S1/S2 cleavage site– and additionally evaluated the impact of E340D. V36M conferred the highest infectivity across all cell lines, with the most significant effect in low-TMPRSS2 cells. While all mutations showed enhanced infectivity with the addition of E340D, the effect was most pronounced in mutations with lower baseline infectivity. The addition of E340D significantly decreased relative neutralizing titres for V36M, S98F, and V213G, enabling escape from neutralizing antibodies in XBB-responsive individuals, illustrating an enhanced phenotypic advantage. Patient neutralizing activity was absent pre-sotrovimab, and sotrovimab-induced neutralization was further compromised by selection of E340D.

**Interpretation:** Sotrovimab pre-exposure prophylaxis in an immunocompromised patient did not prevent SARS-CoV-2 infection, and selected for resistant mutation E340D, with unexpected fitness consequences across non-receptor binding domain spike regions.

**Funding:** Wellcome Grant (220540/Z/20/A), Wellcome Sanger Institute Quinquennial Review 2021

Hong Kong Jockey Club Global Health Institute

NIHR Cambridge Biomedical Research Centre (NIHR203312)

Gates Cambridge Trust

Harding Distinguished Postgraduate Scholars Programme

Glaxosmithkline (GSK)

Xunta de Galicia ED481A-2023/089

## INTRODUCTION

Immunocompromised individuals represent approximately 4% of England’s population and constitute an extremely vulnerable group, disproportionately burdened by severe COVID-19 outcomes. Despite receiving three or more vaccine doses, these individuals account for nearly a quarter of hospitalizations, intensive care unit admittance, and deaths^1^. Suboptimal protection arises from poor immunogenicity following vaccination, characterized by low seroconversion and neutralizing IgG responses^2,3^, resulting in shorter duration of protection, delayed viral clearance, and increased risk of COVID-19 complications^4,5^.

Incomplete viral clearance during chronic SARS-CoV-2 infection in the immunocompromised provides a setting for sustained viral replication and intra-host evolution - conditions theorized to contribute to the emergence of novel variants^6–8^. In this context, the potential use of monoclonal antibodies such as AZD8895/tixagevimab–AZD1061/cilgavimab^9^, AZD3152/sipavibart^10^, and VYD222/pemivibart^11^ as pre-exposure prophylaxis (PrEP) have emerged as an additional preventive strategy to reduce symptomatic SARS-CoV-2 infection within this vulnerable population. However, the evolutionary and clinical implications of breakthrough infections under combined vaccine and monoclonal PrEP remain unclear given the novelty of this intervention.

Treatment-emergent monoclonal antibody resistance has been documented across SARS-CoV-2 lineages, with therapeutic agents differing in their genetic barriers to resistance, as defined by the number and combination of mutations required for viral escape ^12^. In response, immunobridging approaches have been used to accelerate the approval and accessibility of variant-adapted monoclonal antibodies by comparing serum neutralizing titres against emerging variants during a trial with established correlates of protection from monoclonal antibodies targeting earlier variants, as shown for pemivibart^11,13^.

The use of PrEP may influence intra-host SARS-CoV-2 evolutionary dynamics post-infection. PrEP has been documented to interfere with endogenous vaccine-elicited and infection-induced antibodies through epitope masking by shielding antigenic epitopes from relevant B cells^14^. Consequently, the interplay between PrEP exposure, vaccine-elicited antibodies, and viral evolution during active infection warrants investigation to dissect the temporal landscape of resistance emergence, and selection of mutations that may contribute to the development of novel variants. Here, we report the emergence and subsequent selection of VIR-7831/sotrovimab^15^ resistance following PrEP exposure in a haematological patient with chronic SARS-CoV-2 infection, accompanied by a dynamic intra-host mutational landscape and evidence of recombination.

## METHODS

### Clinical report

An immunocompromised male in their 80s with a haematological/oncological diagnosis was enrolled in the PROphylaxis for paTiEnts at risk of COVID-19 infecTion (PROTECT-V) trial (ClinicalTrials.gov NCT04870333; EudraCT 2020-004144-28) sponsored through Cambridge University Hospitals NHS Foundation Trust and the University of Cambridge. The protocol was approved by the UK Medicines and Healthcare Products Regulatory Agency and South Central Berkshire Research Ethics Committee (REC Reference 20/SC/0403). The individual was randomized to receive either a single intravenous infusion of sotrovimab or a placebo, followed by standard of care. Follow-up assessments included telephone and in-person visits such as clinical assessment and blood sample collection. Upon suspected COVID-19 infection, the participant performed a lateral flow test and provided a nasopharyngeal swab for qPCR confirmation. Sampling continued weekly or until two consecutive negative PCR results. Patient sera was collected and stored at -80°C at screening, day 29, day 85, day 169, day 253 after investigational medicinal product administration.

### qPCR-based confirmation and quantification of SARS-CoV-2

Nasopharyngeal swabs were transported in SIGMA VIROCULT^®^ medium (Medical Wire and Equipment Co.) for downstream processing. Viral RNA was inactivated and isolated from samples using QIAamp Viral RNA Mini Kit (QIAGEN, Cat. No. 52904) according to the manufacturer’s protocol. cDNA synthesis occurred in two steps. First, 500 ng of extracted viral RNA was mixed with 1 µL of Oligo(dT)20 Primer (50 μM) (Invitrogen Cat. No. 18418-020), 1 µL of dNTP (10 mM) (Invitrogen Cat. No. 18427-013) and 13 µL of nuclease-free water at 65°C. Second, a mixture 1 µL of RNaseOUT^TM^ Recombinant RNase Inhibitor (Invitrogen Cat. No. 10777019), 1 µL of SuperScript^TM^ III Reverse Transcriptase (Invitrogen Cat. No. 18080093), 4 µL of 5X First-Strand Buffer (Invitrogen Cat. No. 18080093), and 1 µL of 0.1M DTT (Invitrogen Cat. No. 18080093) was added, and the reaction was incubated at 50°C for 60 minutes followed by 70°C for 15 minutes.

cDNA templates were diluted 1:5 in nuclease-free water prior to qPCR. Each reaction contained 50% Fast SYBR Green Master Mix (2X) (Applied Biosystems™. Cat. No. 4385612), forward (GCCTCTTCTCGTTCCTCATCAC) and reverse (AGCAGCATCACCGCCATTG) primers (10 µM each) targeting the SARS-CoV-2 nucleocapsid (N) gene. Relative abundance was quantified by comparing sample Ct values to a standard curve generated from known quantities of a control of the N gene (New England Biolabs. Cat. No. N2117S) included in each qPCR run. Cycling conditions were as follows: initial hold at 95°C for 20 seconds; 40 PCR cycles at 95°C for 1 second, 60°C for 20 seconds; followed by a melt-curve stage of 95°C for 15 seconds, 60°C at 1 minute, and 95°C for 15 seconds.

### Whole-genome sequencing

Whole-genome sequencing occurred using Illumina NovaSeq. Libraries were prepared using the ARTIC V4.1 protocol to generate 400 nucleotide amplicons spanning the viral genome (amplicon) and through bait-capture enrichment method.

### Bioinformatic processing and variant calling

CRAM files were converted to paired FASTQs with SAMtools^16^ and low-quality bases and adaptor sequences were trimmed using Trim Galore (default setting) (https://github.com/FelixKrueger/TrimGalore). Reads were mapped to Wuhan-Hu-1 reference sequence (NC_045512.2) using BWA^17^ and primers were removed with iVar prior to consensus sequence generation at a frequency threshold of 50% with a minimum depth of 10 reads and a minimum base quality score of 20. Variants were called using haplotype-aware FreeBayes^18^ and annotated using bcftools csq^19^ using a modified Wuhan-Hu-1 GenBank file to include ORF3c^20^ and ORF9b^21^, in localized consequence-aware mode – optimized for short-read, multiallelic data – at a frequency threshold of 2% with a minimum depth of 10 reads, minimum base quality score of 20, and minimum base mapping quality score of 20. Low-frequency mutations were defined as 2-49%. Pipeline details are available at https://github.com/TKMarkCheng/ProtectV_seq.

### Alignment and masking

Sequences were aligned using MAFFT (v7.525) with the auto, reorder, anysymbol and keeplength parameters^22^. Known position exhibiting suspected sequencing artifacts or high ambiguity were masked prior to analysis (https://github.com/W-L/ProblematicSites_SARS-CoV2, commit: a36cee5).

### Recombination detection

Screening for recombination is standard practice prior to phylogenetic reconstruction as recombination violates the assumption that the viral genealogy can be accurately modelled as a single bifurcating tree. To detect any possible recombination events, we used Genetic Algorithm Recombination Detection (GARD) to scan sequence alignment and identify potential breakpoint locations^23^. Segment-specific phylogenetic trees were produced from non-recombinant regions and breakpoint support was evaluated using Akaike’s Information Criterion (AIC). GARD evaluates multiple models with different numbers and placements of breakpoint(s). For each potential breakpoint placement, AIC is computed and the average is outputted as model-average support.

Ancestral recombination graphs (ARGs) were constructed over non-recombinant regions by removing the recombinant region associated with low model support using the Espalier package through Python. Espalier computes maximum agreement forests (MAFs) between discordant phylogenetic trees by calculating the fewest number of branch cuts required to achieve topological concordance through a prune-and-regraft approach and visualizes the resulting, if any, discordance using tanglegrams.

### Phylogenetic analyses

We performed phylogenetic analyses of the consensus level genomes from the patient using BEAST 2^24–31^. Given the putative recombination identified above, we used bacter^32^, an implementation of the ClonalOrigin model^33^, for our tree generating stochastic process. This modification to the coalescent allows one-way donation of genomic segments across the tree, allowing recombination events to be explicitly modelled. The precise specification of all phylogenetic models and priors used can be found in nSupplementary Table 1a. Sensitivity analysis was performed on the parameter controlling the rate of gene conversion, as this could not be informed from the limited data. The reported substitution rate comes from a model with an expected rate of gene conversions of 0.0001 conversions/year.

High coverage GISAID sequences with collection dates between May 2023 to February 2024 were downloaded (n=248, EPI_SET_250930sz; Supplementary Table 1b) and merged with the first bait-capture sequence from the focal patient. All sequences were aligned and processed, including masking, as described above. Phylogenetic inference was performed using IQ-TREE (v3.0.1) under the GTR+I+R_4_ substitution model. Temporal reconstruction was carried out with TreeTime (v0.11.4) on the final tree rooted to Wuhan-Hu-1 (NC_045512.2) and visualized using R (v4.5.1)^34,35^. The resultant tree was used as an initialisation condition for Bayesian analyses in BEAST 2 using metropolis-coupled MCMC to assess tree uncertainty and estimate substitution rates.

Following detection of the potential recombination event, participant sequences were divided into two regions representing non-recombinant segments, and independent trees with linked parameters were built for each region in BEAST 2.

For all Bayesian phylogenetic analyses, convergence was assessed by running four independent runs and comparing between them using standard approaches. The first 50% of each independent run was discarded as burn-in. In all cases, consensus sequences or partial consensuses were assumed to be instantiated in real virions within the patient, making the above a coherent approach for estimating the virion-level substitution rate - the standard phylogenetic estimand, which is a complex product of selection and mutation.

### Selection analysis and mutations chosen for downstream functional characterization

In part, we used the codon-based evolutionary model Fast, Unconstrained Bayesian AppRoximation (FUBAR) method^36^ to identify codon sites under positive or purifying selection within the spike (S) gene.

Considering S mutations with dynamic temporal profiles and variable within-host read prevalence, we prioritized five nonsynonymous mutations for further functional characterization. Three were located in the N-terminal domain (V36M, S98F, V213G), one in the receptor-binding domain (Y505P), and one upstream of the polybasic S1/S2 cleavage site (P681Q). S98F and V213G were identified at consensus thresholds through both amplicon and bait-capture methods while V36M, Y505P, and P681Q were identified at low-frequencies solely through the amplicon method.

### Site-directed mutagenesis

Plasmids encoding the SARS-CoV-2 S protein were used for pseudotyped virus production and kindly provided by J. Newman (Pirbright Institute), T. Peacock and W. Barclay (Imperial College London, UK) via the Genotype-to-Phenotype National Virology Consortium (G2P- UK). S mutations were generated using pcDNA3.1 plasmids that were codon-optimized and included a Δ19 deletion at the C terminus of the GE.1 S gene. The GE.1 backbone included mutations T19I, L24S/Δ25/27, V83A, G142D, Δ144, H146Q, Q183E, Δ185/F186I, V213E, D253G, G339H, R346T, L368I, S371F, S373P, S375F, T376A, D405N, R408S, K417N, N440K, V445P, G446S, N460K, S477N, T478R, E484A, F486P, F490S, Q498R, N501Y, Y505H, P521S, D614G, H655Y, N679K, P681H, N764K, D796Y, Q954H, N969K. Additional mutations including V36M, S98F, V213G, Y505P, and P681Q were introduced to assess functionality of novel S mutations and were incorporated into the plasmid backbone using QuikChange II Site-Directed Mutagenesis Kit (Agilent Cat. No. 200523).

### Cell culture and maintenance

HEK293T cells were used as pseudotyped virus producer cells. HeLa-ACE2 were used as target cells to examine sera neutralizing antibodies against pseudotyped virus entry in neutralization assays. HeLa-ACE2, A549-ACE2/TMPRSS2, A549-ACE2, Caco-2, and Calu-3 cell lines were used to examine viral entry in infectivity assays. HEK293T, HeLa-ACE2, A549-ACE2/TMPRSS2, and A549-ACE2 cells were cultured in Dulbecco’s Modified Eagle Medium (DMEM) (ThermoFisher Scientific Cat. No. 11965092) supplemented with glutamine, 10% foetal bovine serum (FBS) (Sigma-Aldrich Cat. No. F7524), 1% penicillin-streptomycin (100 U/mL penicillin and 0.1 mg/mL streptomycin) (ThermoFisher Scientific Cat. No. 15140122) at 37°C with 5% CO_2_. Caco-2 cells were maintained in DMEM with 10% FBS, 1% non-essential amino acid (NEAA) (ThermoFisher Scientific Cat. No. 11140050), 1% Sodium Pyruvate (ThermoFisher Scientific Cat. No. 11360070), 1% penicillin-streptomycin (100 U/mL penicillin and 0.1 mg/mL streptomycin) at 37°C with 5% CO_2_. Calu-3 cells were maintained in Minimum Essential Medium (ThermoFisher Scientific Cat. No. 21090022) with 10% FBS, 1% NEAA, 1% sodium pyruvate, 1% GlutaMAX^tm^ (ThermoFisher Scientific Cat. No. 35050061), 1% penicillin-streptomycin (100 U/mL penicillin and 0.1 mg/mL streptomycin) at 37°C with 5% CO_2_. Vero E6 were maintained in DMEM with 10% FBS, G418 mg/mL (Merck Cat. No. G8168), and 1% penicillin-streptomycin (100 U/mL penicillin and 0.1 mg/mL streptomycin) at 37°C with 5% CO_2_.

### SARS-CoV-2 pseudotyped virus production and neutralization assay

The methodological details have been reported previously by the authors^3^. Briefly, viral entry of GE.1 SARS-CoV-2 harbouring different mutations in the presence of patient neutralizing sera was quantified using luciferase-expressing lentiviral pseudotyped virus. Samples were thawed, heat-inactivated at 55°C for one hour, and diluted 1:10. Diluted serum was incubated with pseudotyped viruses and HeLa-ACE2 cells, followed by incubation for 48 hours at 37°C with 5% CO_2_, after which luminescence was measured as the relative light unit (RLU) using the Bright-Glo Luciferase Assay System (Promega). Data were normalized relative to pseudotyped virus-only and cell-only controls. All neutralizing assays were repeated in two independent experiments, each comprising two technical replicates per condition.

We evaluated neutralizing titres against pseudotyped viruses containing novel mutations using two approaches. First, we assessed the impact of these mutations in immunocompetent individuals by analysing sera from individuals from the NBR118 cohort with detectable XBB-neutralizing titres post-dose 3 and post-dose 4 vaccination^3^, to determine whether mutations reduced neutralization in responsive individuals. The NBR118 study was approved by the East of England Cambridge Central Research Ethics Committee (17/EE/0025). Second, we measured longitudinal samples from the clinical patient collected throughout their chronic infection including screening, 1-and 29-days post-infusion of sotrovimab, 28 days post-dose 7 vaccination, 90-days post-infusion of sotrovimab, day 1 of SARS-CoV-2 infection, 28-days post infection, and 28-days post-dose 8 vaccination.

### Quantification of reverse transcriptase activity as a measure of viral infectivity

A modified SYBR Green I-based qPCR product-enhanced reverse transcriptase (SG-PERT) assay has previously been used to measure reverse transcriptase activity in lentiviral vectors^37^. SG-PERT measurements correlate with the number of transducing pseudotyped viral units. In parallel, pseudotyped virus was incubated with different target cell lines. Together, SG-PERT values and luminescence measured using the Bright-Glo Luciferase Assay System (Promega) served as complementary surrogate measures of cellular infectivity, accounting for viral input. In detail, pseudotyped virus was added to an equal volume of lysis solution (40% glycerol, 0.25% Triton X-100, 50mM KCl, 1M TrisHCl pH7.4, 800 U/mL of Ribolock RNase inhibitor; Thermo Scientific™ Cat. No. EO0381) and incubated for 10 minutes at room temperature. Viral lysates were then added at a 2:3 ratio to a reaction mix containing 83% QuantiTect SYBR Green PCR Master Mix (QIAGEN Cat. No. 204143), 0.5 uM forward MS2 primer (TCCTGCTCAACTTCCTGTCGAG), 0.5 uM reverse MS2 primer (CACAGGTCAAACCTCCTAGGAATG), 500 U/mL Ribolock RNase inhibitor (Thermo Scientific™ Cat. No. EO0381), and 3.5 pmol/mL MS2 RNA (Roche Cat. No. 10165948001). Standards of known activity (HIV reverse transcriptase, recombinant, *E.coli*; Millipore Cat. No. 382129) were included in each run. Reactions were performed using the QuantStudio 7 Flex Real-Time PCR System, and reverse transcriptase activity for each pseudotyped virus was determined from the rate of MS2 RNA amplification relative to the standard curve generated from the recombinant HIV reverse transcriptase. Cycling conditions were as follows: initial hold at 42°C for 20 minutes and 95°C for 15 minutes; 40 PCR cycles at 95°C for 10 seconds, 60°C for 30 seconds, and 72°C for 15 seconds; followed by a melt-curve stage of 95°C for 15 seconds, 60°C at 1 minute, and 95°C for 15 seconds.

Pseudotyped viruses were then used to transduce target cell lines (HeLa-ACE2, A549-ACE2/TMPRSS2, A549-ACE2, Caco-2, and Calu-3) in a 1:1 ratio and were incubated for 48 hours at 37°C with 5% CO_2_. Luminescence (RLU) was measured using the Bright-Glo Luciferase Assay System (Promega), and data were normalized to SG-PERT-derived values to report RLU per unit of reverse transcriptase activity (RLU/U).

### Cell-cell fusion assay

GFP1-10 expressing HEK293T cells and Vero E6 cells stably expressing GFP11 were mixed 1:1 ratio at 80% confluence and co-transfected with 250 ng GE.1 plasmids containing V36M, S98F, V213G, Y505P, P681Q, V36M+E340D, S98F+E340D, V213G+E340D, Y505P+E340D, P681Q+E340D, or GE.1+E340D using FuGENE 6 Transfection Reagent (Promega Cat. No. E2691). Fusion of cells resulted in detection of GFP signal and was determined as the proportion of green area to the total phase area. Imaging and analysis examining cell-cell fusion occurred at 0-, 6-, 12- and 18-hours post-transfection using the IncuCyte SX5 Live-Cell Analysis System from Sartorius.

### Statistical analysis

Neutralizing IC_50_ were calculated using GraphPad Prism version 10.6.1. Assay limits of detection ranged between 40 and 87480, and values were constrained to these limits. Neutralizing values were summarized using geometric mean titre (GMT) with geometric standard deviation. Infectivity was reported as RLU per unit of reverse transcriptase activity (RLU/U) based on each mutant. Data analysis was performed using statistical software R. The significance is annotated as *p < 0.05; **p < 0.01; ***p < 0.001; ****p < 0.0001.

## RESULTS

### Case information

An immunocompromised man in their 80s with chronic lymphocytic leukaemia enrolled in the PROTECT-V trial. The individual was enrolled and screened in March 2023 and randomized to receive a single 2000 mg intravenous infusion of sotrovimab in 03-2023. At time of enrolment, the individual had received six SARS-CoV-2 vaccinations (Figure 1A) and subsequently received two additional boosters by the end of the study period. Anti-spike IgG levels at screening were <0.4 U/mL, increased to 18250 U/mL prior to a positive lateral flow test in 08-2023, and subsequently decreased during the infection period. The participant was confirmed positive by qPCR on 08-2023, providing weekly swabs (n=24) from 08-2023 to 02- 2024 (171 days) (Figure 1). All samples were confirmed positive by qPCR and had viral loads ranging from 1.4e+06 to 8.4e+13 RNA copies/mL.

**Figure 1.**
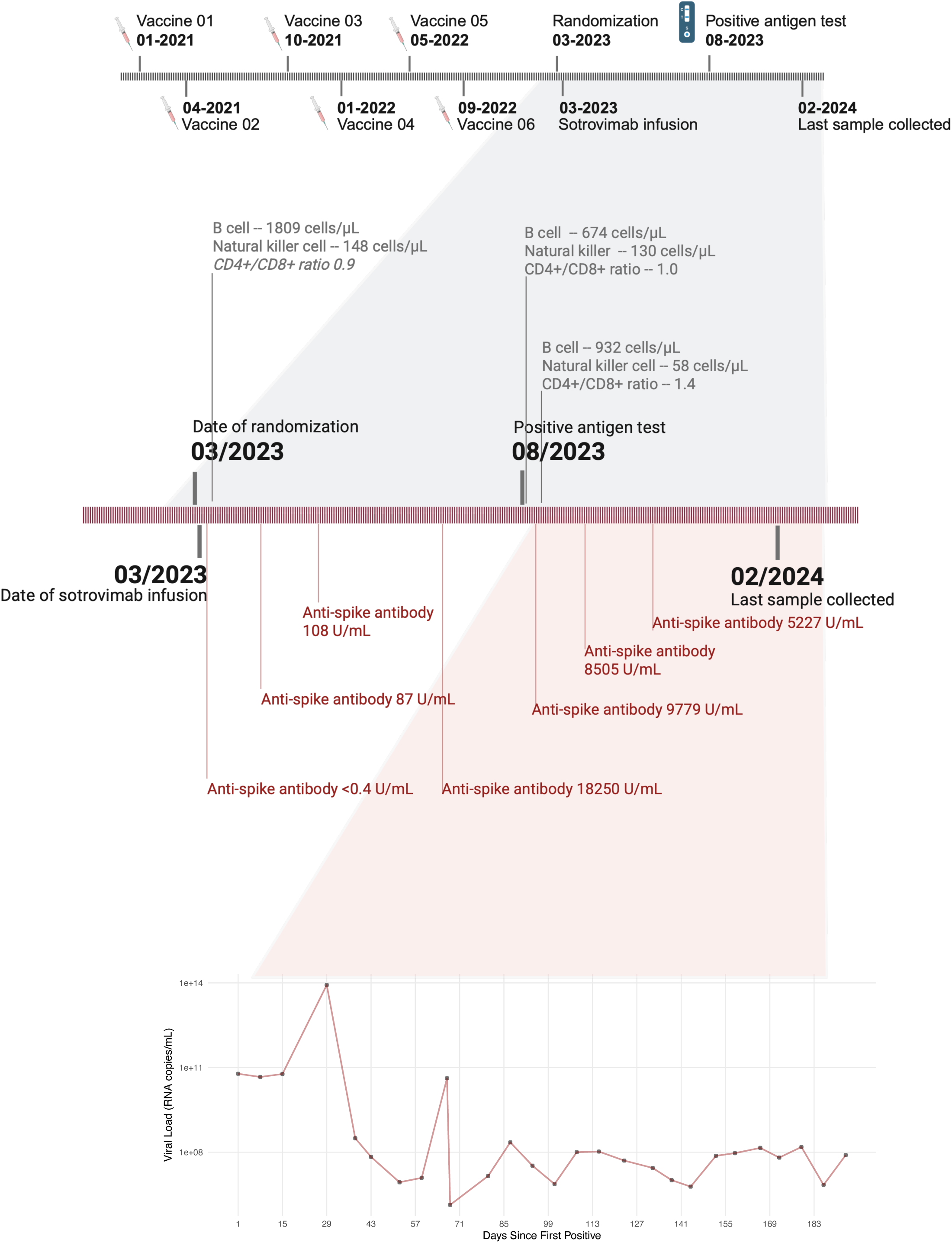
Timeline of sotrovimab treatment, SARS-CoV-2 infection, and viral load measurements. The participant received six SARS-CoV-2 vaccinations prior to Sotrovimab infusion in 03- 2023. Anti-spike antibody levels were <0.4 U/mL at time of screening and increased following sotrovimab administration, remaining detectable throughout the infection period. The individual first tested positive for SARS-CoV-2 through a lateral flow test in 08-2023, with infection confirmed by qPCR a week after. Weekly nasopharyngeal swabs were collected from 08-2023 to 02-2024 (n=24; 171 days). All samples tested positive by qPCR, with viral loads ranging from 1.4e+06 to 8.4e+13 RNA copies/mL, as semi-quantified through qPCR.

### Viral sequencing and mutations

All 24 samples were successfully sequenced using both amplicon and bait-capture methods and were classified as the GE.1 variant by CoV-lineages PANGOLIN v4.3.1. Amplicon sequencing achieved a median genome coverage of 97.0% (Q1-Q3: 96.0–97.3%), while bait-capture based sequencing achieved 98.2% (Q1-Q3: 98.2–98.3%). After accounting for lineage-defining mutations across the infection period, matrix (M) gene had the highest mutation density per kilobase (kb) across both sequencing methods (amplicon 105, bait-capture 61; Supplementary Figure 1A) although ORF1ab and the S gene had the highest overall counts of mutations (amplicon 447, bait-capture 339; amplicon 228, bait-capture 175, respectively; Supplementary Table 2). Amplicon sequencing detected a higher number of low-frequency (2-49%) mutations during infection, whereas bait-capture sequencing exhibited greater consistency in the mutations identified, including lineage-defining mutations (Supplementary Figure 1B). These findings align with previous reports indicating that both amplicon and bait-capture methods provide concordant sequencing, with bait-capture offering higher uniformity in coverage and amplicon sequencing exhibiting greater sensitivity for detecting low-frequency mutations^38^. Accordingly, we used bait-capture-derived data for analyses requiring consensus sequences and amplicon-derived data for low-frequency analysis.

### Temporal shift in mutation profiles in S gene and signatures of recombination in ORF1ab

S98F and E340D mutations in the S gene were consistently detected at high abundance throughout the infection (Figure 2A). S98F remained fixed at 100% frequency across all timepoints (24 weeks; 171 days), while E340D – a known sotrovimab-resistance mutation – fluctuated over time and ultimately reached a frequency of 96% by week 15 (107 days post-infection) (Figure 2B, Figure 2C). Between week 14 and week 15 (100- and 107-days post-infection), we observed a marked shift in the low-frequency mutation landscape (Figure 2A; Supplementary Figure 1C). This shift coincided with a 14-fold increase in viral load (7.42e+06 to 1.00e+08 RNA copies/mL). Although not the highest recorded viral load of this patient, this rebound following a period of decline together with the change in mutation profile prompted further investigation.

**Figure 2.**
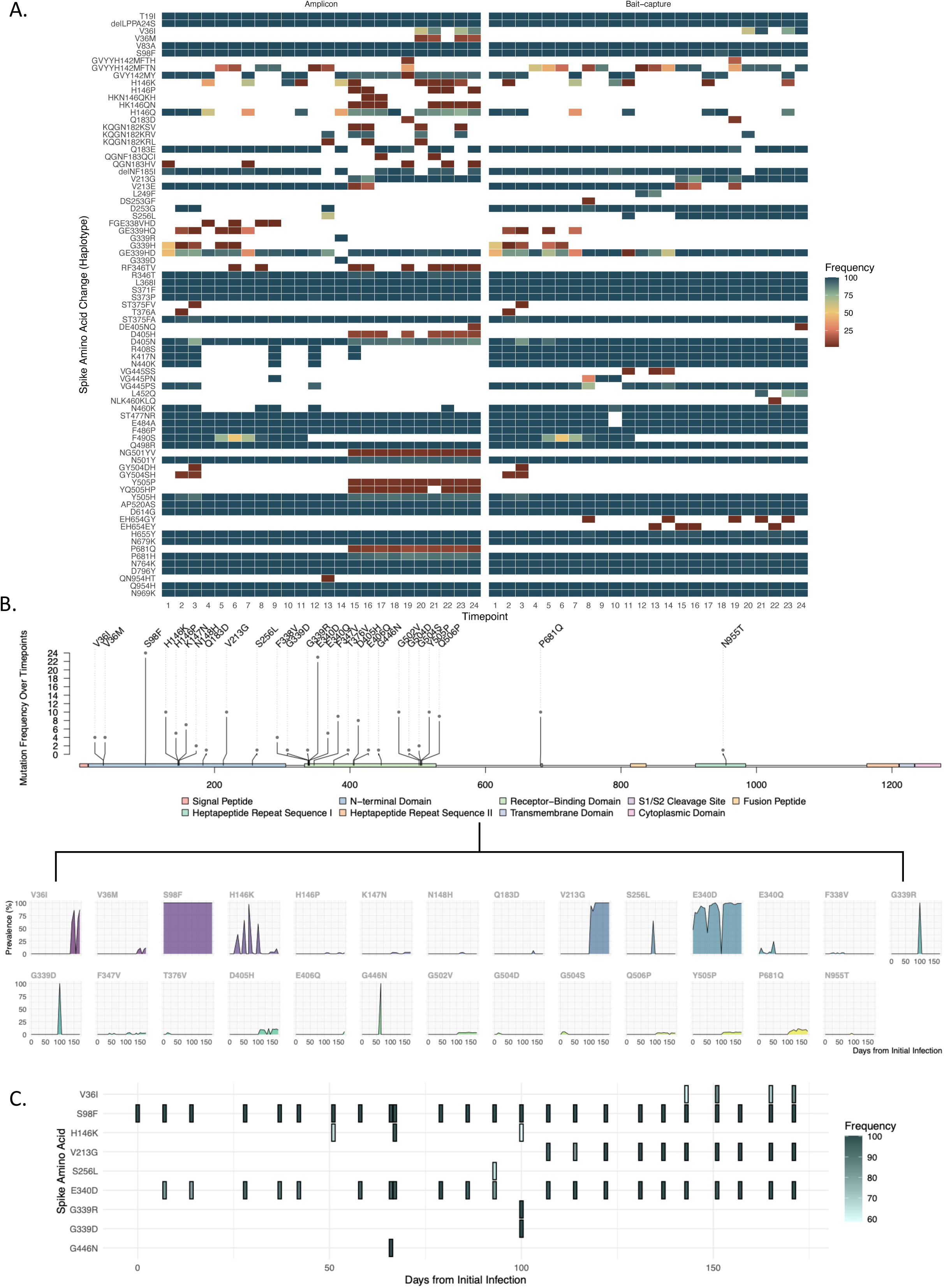
Temporal changes in mutation spectrum revealed the emergence and selection of novel mutations in the spike gene. A) Haplotype amino acid analysis showed combinations of lineage and non-lineage mutations. S98F and E340D were consistently abundant, with E340D co-occurring with the Omicron-defining G339H. B) Counts and prevalence of non-lineage defining mutations highlight repeated changes at the same amino acid positions (e.g., V36I vs V36M; G339R vs G339D). E340D, initially present at non-consensus level, fluctuated before being selected alongside V213G around 100 days post-infection. C) Consensus-level non-lineage defining mutations consistently present included S98F and E340D, with V213G joining 107 days post-infection.

We detected a recombination breakpoint within the ORF1ab gene spanning positions between 8942 to 12458 (Figure 3A, Supplementary Figure 2). The sequences obtained from week 23 and week 24 (165- and 171-days post-infection) exhibited the putative recombination event implying that the recombination event must have occurred prior to 165 days post-infection. The region of recombinant genomes 5’ of the breakpoint most closely resembled sequences sampled from the early part of the infection (weeks 2-10, 12-14), whereas the region 3’ of the breakpoint most closely resembled the viral population sampled after week 14. Figure 3B illustrates the mosaic phylogenetic histories of the two genomic regions by reconstructing ancestral recombination graphs (ARGs) using maximum agreement forests (MAFs). The recombination event is marked by sample week 24 falling in different clades between the tree from before and after the putative breakpoint.

**Figure 3.**
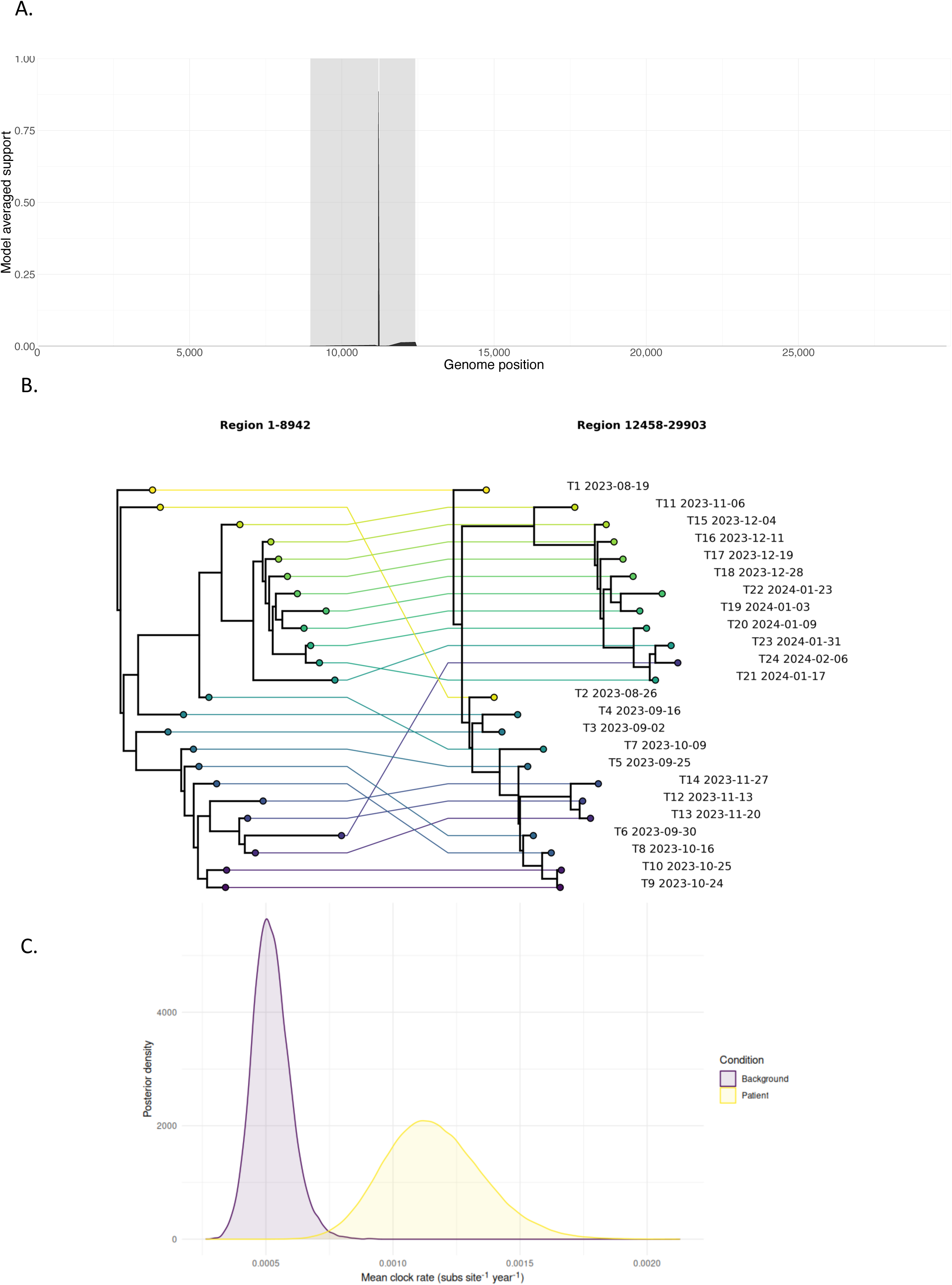
Recombination analysis identified potential breakpoint in the ORF1ab region. A) GARD analysis identified phylogenetic incongruence across the alignment with a recombination breakpoint between positions 8942 and 12458. Uncertainty in the exact breakpoint location is reflected in the variation in the breakpoint placement support. Grey shading indicates regions of increasing model-averaged support for breakpoint placement, while black shading represents the model-averaged support values within that region, peaking at the most strongly supported position. B) A tanglegram constructed from genomic regions prior to and after the recombination breakpoint. The cladograms represent the two non-recombinant genome segments with linkages inferred using maximum agreement forests and indicating the discordance between the topologies. C) Posterior distribution from the clock rate for a tree built from all GE.1 sequences on GISAID and the first sample from the patient (Background) under the GTR+R4 model with a lognormal relaxed clock and skyline tree prior and one built from the patient (Patient) under the GTR+R4 model with a strict clock and bacter tree prior.

### Intra-host and inter-host virion-level substitution rates

Phylogenetic models allowed the calculation of the intra-host virion-level substitution rate (substitutions/site/year) for this patient from their 24 longitudinal samples, and the GE.1 inter-host rate from 249 global GE.1 sequences acquired from GISAID. All sequences from the patient clustered together, suggesting that all within-patient diversity was associated with a single infection event (Supplementary Figure 3). The estimated intra-host rate was 1.13e-3 substitutions/site/year (95% highest posterior density interval: 7.54e-4 - 1.51e-3), compared to the GE.1 inter-host rate of 5.22e-4 substitutions/site/year (95% highest posterior density interval: 3.79e-4 - 6.72e-4) (Figure 3C), with direction of this effect being robust to sensitivity checks (Supplementary Figure 4).

### Site-specific selection in spike and assessing functionality

The ratio of nonsynonymous substitution rate (dN) to synonymous substitution rate (dS) (ω) is one of the main ways site-specific diversity is assessed to infer neutral, positive, or negative selection. Sites 36, 146, 213, 446, 461, and 490 were identified as sites with high ω indicative of positive selection (Supplementary Figure 5A; Supplementary Table 3). Two sites, 335 and 541, were identified under negative selection (Supplementary Figure 5B). Looking at mutations more closely, four sites (146, 213, 446, and 490) were Omicron-defining positions and three sites (446, 461, and 490) were within the ACE2 binding region.

We prioritized the functional assessment of five nonsynonymous mutations in the S gene: three in the N-terminal domain (V36M, S98F, V213G), one in the receptor-binding domain (Y505P), and one upstream of the polybasic S1/S2 cleavage site (P681Q). These mutations encompassed both consensus-level (S98F and V213G) and low-frequency (V36M, Y505P, and P681Q) mutations. Our analysis focused on lineage-defining positions (213, 505, and 681) that harboured novel amino-acids changes, as well as fixed mutations such as S98F.

We also evaluated the impact of E340D, a sotrovimab resistance mutation, in combination with each mutation to assess potential epistatic effects. Although E340D was present at a frequency of 46% at week 1 of infection, its frequency fluctuated during the first 13 weeks, before reaching 96% by week 15 (107 days post-infection).

### Increased cell infectivity with acquisition of E340D

The addition of E340D to the GE.1 backbone had modest but statistically significant effect on cellular entry in HeLa-A2, Caco-2, and A549-A2T2 cells (Lognormal Welch’s t test with Bonferroni-Dunn adjustment, p-value<0.05 for all; Supplementary Figure 6A). Mutation Y505P yielded limited detectable luciferase signal and was omitted from further analyses (Supplementary Figure 6B).

V36M increased infectivity in HeLa-A2, Calu-3, and A549-A2 cells (one-way ANOVA with Dunnett’s multiple comparisons test, p-value<0.05 for all), although it was most apparent in low TMPRSS2/high-ACE2 expressing cell lines (HeLa-A2 and A549-A2) (Figure 4A). While all mutations showed enhanced infectivity with the addition of E340D, the effect was most pronounced in mutations with lower baseline infectivity (Figure 4B). For example, although V36M exhibited the highest infectivity across all cell types, its fold change with E340D was smaller compared to other mutations (log2 fold-change: 0.9 in HeLa-A2, 1.1 in Calu-3, 3.0 in Caco-2, 3.2 in A549-A2T2, and 4.6 in A549-A2) (Supplementary Table 4). Overall, the GE.1 backbone plus E340D displayed the greatest enhancement relative to the GE.1 backbone, ranging from 1.5-fold in Calu-3 cells to 6-fold in A549-A2 cells (Figure 4B).

**Figure 4.**
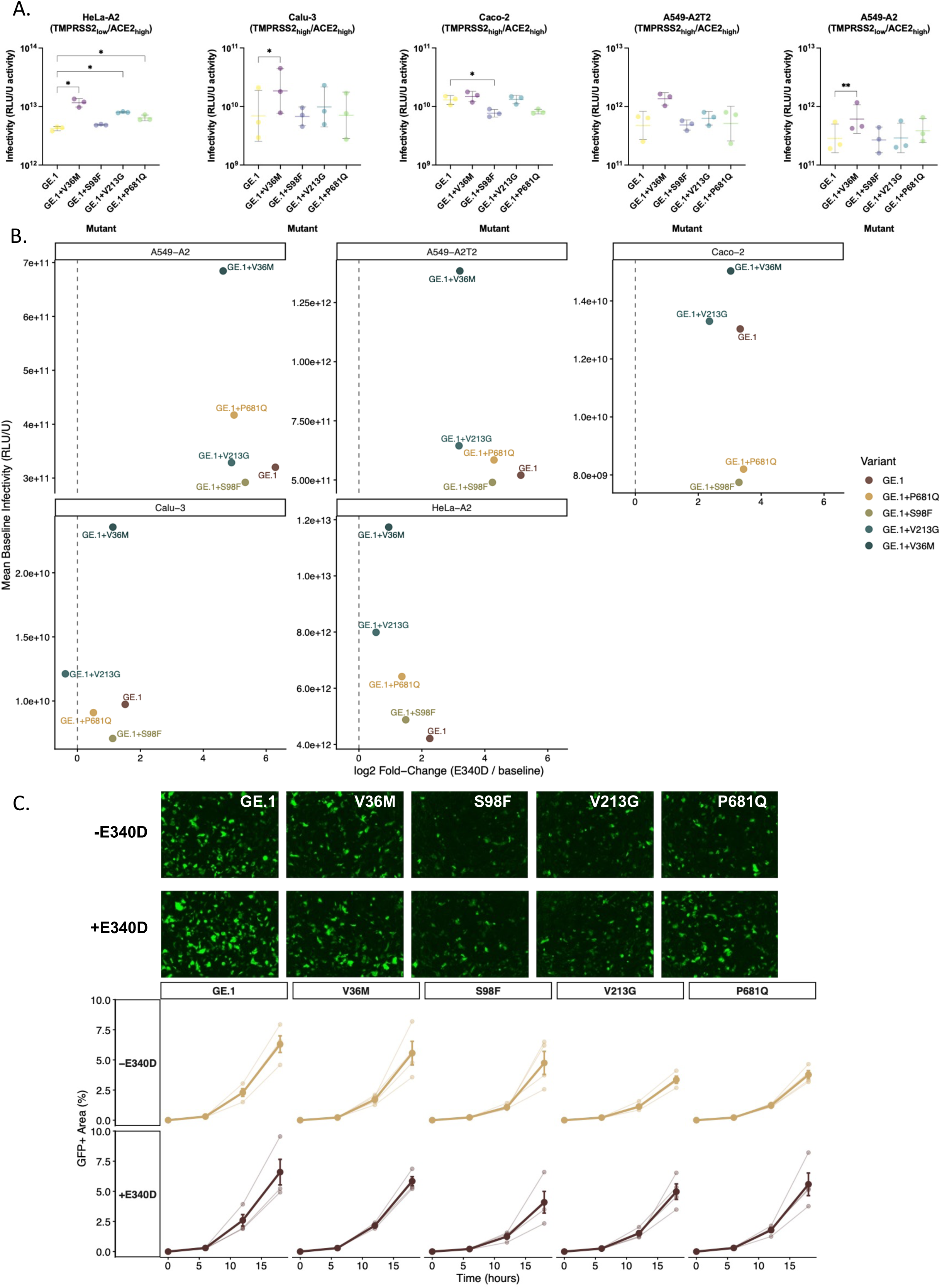
Spike mutations indicative of altered Omicron tropism and epistatic effects of E340D on cellular infectivity in combination with other spike mutations. A) V36M increased infectivity relative to the backbone in HeLa-A2, Calu-3, and A549-A2 cells (one-way ANOVA with Dunnett’s multiple-comparisons test against backbone; adjusted p-value=0.025, 0.037, and 0.0021, respectively). In contrast, S98F decreased infectivity relative to the backbone in Caco-2 cells (adjusted p-value=0.011), consistent with Omicron’s shift from gastrointestinal to respiratory tropism. B) Most spike mutations exhibited increased infectivity with the addition of E340D, with the largest enhancements observed among mutations with lower baseline infectivity. V36M, despite the highest baseline infectivity, exhibited smaller fold changes across cell lines (log2 fold-change: 0.9–4.6). S98F showed enhancement ranging from 1.12-fold to 5.3-fold in A549-A2 cells, while V213G displayed variable effects across cell types, from reduced infectivity in Calu-3 cells (−0.4-fold) to increased infectivity in A549-A2 cells (4.9-fold). P681Q combined with E340D resulted in enhancements ranging from 0.5-fold in Calu-3 to 4.97-fold in A549-A2 cells. The GE.1 backbone plus E340D showed the strongest overall enhancement, ranging from 1.5-fold in Calu-3 to 6-fold in A549-A2 cells. C) Representative images of spike-mediated cell-cell fusion in V36M, V36M+E340D, S98F, S98F+E340D, V213G, V213G+E340D, P681Q, P681Q+E340D, GE.1 and GE.1+E340D at 18 hours post-transfection. Images were captured using the IncuCyte SX5 Live-Cell Analysis System (Sartorius). Quantification of spike-mediated cell-cell fusion showing the percentage of GFP^+^ area to the total cell area at 0-, 6-, 12-, and 18-hours post-transfection. Cell-cell fusion was determined as the proportion of green area to the total phase area and was calculated with the IncuCyte analysis software (Sartorius). Data are mean ± s.e.m. from four fields of views at each time point and are representative of two independent experiments.

V36M, S98F, V213G, and P681Q all reduced syncytium formation relative to the GE.1 backbone, indicated by lower relative GFP^+^ area at 18-hours post transfection (GE.1>V36M>S98F>V213G>P681Q) (Figure 4C). The addition of E340D enhanced cell-cell fusion in all mutants except S98F but remained comparable in the case of the parental backbone (fold change range 0.9-1.5: V36M 1.1, S98F 0.9, V213G 1.5, P681Q 1.5, GE.1 1.0).

### E340D reduced neutralization sensitivity of novel participant mutations in XBB-responsive individuals

Sera from eight individuals from the NBR118 trial (median age 70.5, Q1-Q3:61-73; 50% male) (Supplementary Table 5) with prior detectable XBB-neutralizing titres at post-dose 3 and post-dose 4 COVID-19 vaccination were used to assess whether V36M, S98F, V213G, or P681Q – and their combination with E340D – were associated with decreased neutralization. V36M, S98F, and V213G were associated with increased neutralizing titres and therefore higher susceptibility to neutralizing antibodies relative to GE.1 backbone, (one-way ANOVA with Dunnett’s multiple comparisons test: V36M, p-value=0.0009; S98F, p-value=0.0001; V213G, p-value=0.005) (Figure 5A). The addition of E340D to each mutation significantly decreased neutralizing titres for V36M, S98F, and V213G, indicating that E340D facilitated partial escape from neutralizing antibodies (one-way ANOVA with Šídák’s multiple comparisons test: V36M vs. V36M+E340D, p-value=0.0004; S98F vs. S98F+E340D, p-value=0.001; V213Gvs.

**Figure 5.**
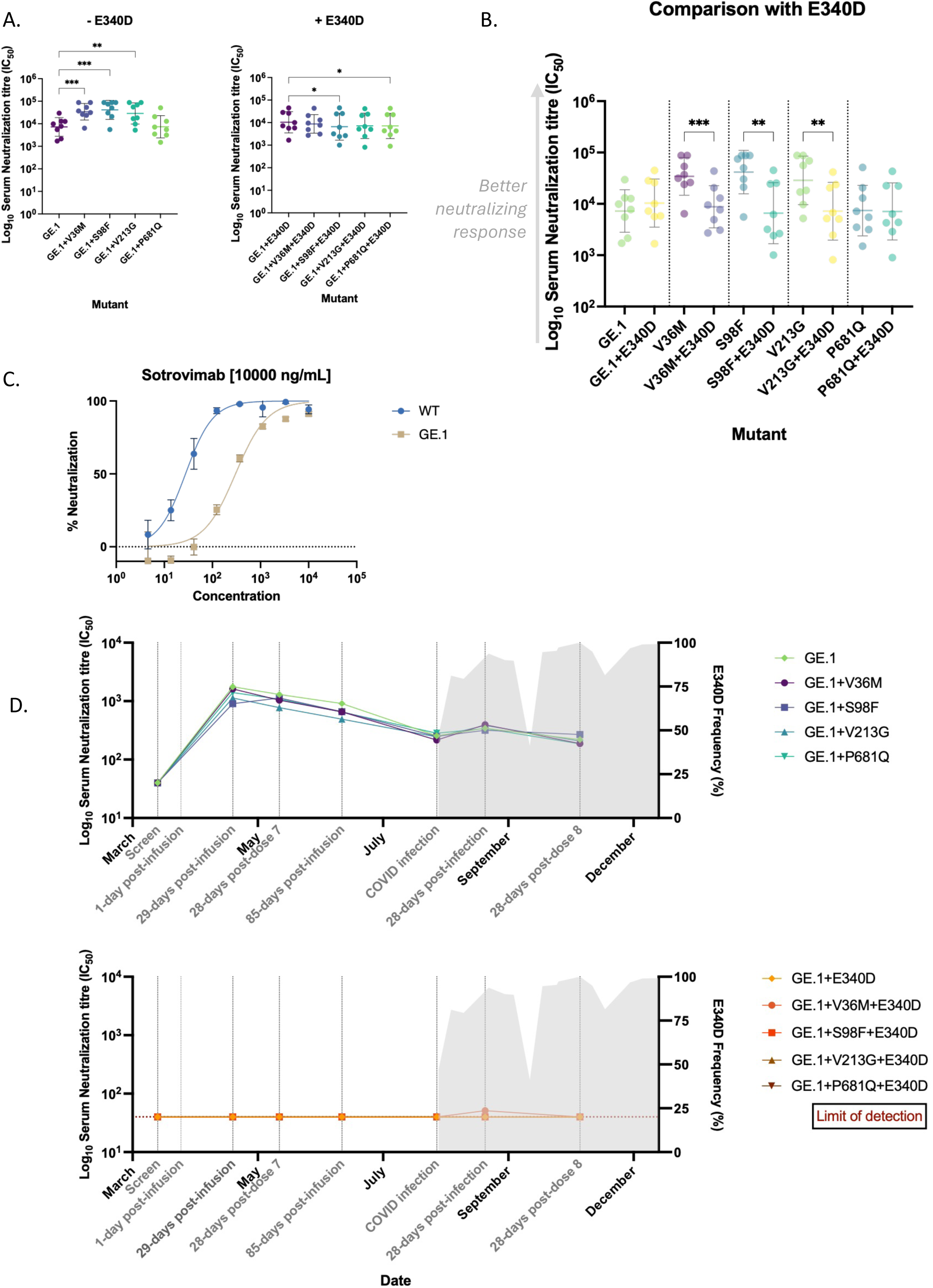
E340D alone and combined with novel mutations, escaped neutralization in XBB-responsive individuals. A) V36M, S98F, V213G, and P681Q exhibited increased neutralization relative to GE.1 backbone, although P681Q did not reach statistical significance (one-way ANOVA with Dunnett’s multiple comparisons test against backbone: V36M, p-value=0.0009; S98F, p-value=0.0001; V213G, p-value=0.005, P681Q, p-value>0.05). Addition of E340D reversed this neutralizing advantage for S98F and P681Q, resulting in reduced neutralization relative to the GE.1+E340D backbone (one-way ANOVA with Dunnett’s multiple comparisons test against GE.1+E340D: S98F, p-value=0.035; P681Q, p-value=0.02). B) Incorporation of E340D significantly reduced neutralization for V36M, S98F, and V213G, consistent with immune-escape from neutralizing antibodies (one-way ANOVA with Šídák’s multiple comparisons test: V36M vs. V36M+E340D, p-value=0.0004; S98F vs. S98F+E340D, p-value=0.001; V213Gvs. V213G+E340D, p-value=0.002). C) Sotrovimab neutralized WT+D614G and GE.1 pseudo virus within the expected concentration range, with the inhibitory concentration (IC_50_) values of 27.6 ng/mL (95% CI: 23.1-33.0 ng/mL) and 302.3 ng/mL (95% CI: 217.3-429.4 ng/mL, respectively. D) At screening, the participant had no detectable neutralizing antibodies against GE.1, or the GE.1 variant carrying mutations V36M, S98F, V213G, P681Q. At 29-days post-sotrovimab infusion, geometric mean titres (GMTs) across all mutations increased 33-fold (from 40 to 1325, SD: 1-1.31). Neutralizing responses subsequently waned, with GMT declining to 248 (SD: 1.1) at time of COVID infection. This represents an approximately 5-fold reduction from post-infusion levels. A slight increase was observed 28-days post-infection (GMT 355, SD:1.1) followed again by waning. Individual mutations did not alter this overall trajectory, as GMTs were comparable across variants 28-days post-infusion (GE.1: 299.0, SD: 4.3; V36M: 269.8, SD: 4.0; S98F: 262.9, SD: 3.6; V213G: 246.0, SD: 3.5; P681Q: 269.9, SD: 3.89). The addition of E340D abolished any response in neutralizing ability that may have been present post-infection, consistent with antibody escape and suggests minimal contribution from host-derived antibodies thereafter.

V213G+E340D, p-value=0.002) (Figure 5B). Interestingly, the addition of E340D in the GE.1 backbone alone slightly though not significantly, increased neutralizing titres (p-value>0.05), suggesting a context-dependent epistatic effect modulated by the presence of other mutations. The inhibitory concentration (IC_50_) of sotrovimab required to achieve 50% neutralization of wildtype+D614G (WT) and GE.1 was 27.6.2 ng/mL (95% CI: 23.1-33.0 ng/mL) and 302.3 ng/mL (95% CI: 217.3-429.4 ng/mL), respectively (Figure 5C). GE.1 is a descendant of the XBB.2 lineage, itself derived from recombinants BM.1 and BJ.1 of the BA.2 lineage. Our findings are within range with previous findings as BA.2 and its progeny typically require approximately 100-1000 ng/mL concentrations of sotrovimab for comparable neutralization^39^. Our results suggest that GE.1 retains sensitivity to sotrovimab within the range expected for a BA.2 descendant.

Patient neutralizing activity was absent pre-sotrovimab, increased post-sotrovimab infusion and became impaired by the presence of E340D (Figure 5D). Based on neutralizing measurements of patient sera containing sotrovimab, geometric mean neutralizing titres 29-days post-sotrovimab infusion were approximately 33-fold higher against the V36M, S98F, V213G, P681Q, and the backbone itself (40 to 1325, SD: 1-1.31) than pre-infusion. However, at time of infection, geometric mean neutralizing titres were only approximately 6-fold higher than pre-sotrovimab levels (40 vs 248, SD: 1-1.1), indicative of waning neutralizing responses. Waning sotrovimab-induced neutralization was further compromised by the early emergence of the E340D mutation. Although E340D was not present at consensus level at week 1, its relative frequency steadily increased over the course of infection and ultimately became fixed. Notably, a similar rise in neutralizing titres was not reproduced at later timepoints despite two additional vaccinations (dose 7 and dose 8) and instead titres continually declined thereafter, with only a brief increase observed post-infection.

## DISCUSSION

In individuals with humoral deficiencies, such as the clinical case presented here, prophylactic monoclonal antibodies are a logical strategy, providing proxy immunity where endogenous responses are lacking to help prevent the risks linked with infection. Yet the resistance threshold for sotrovimab may be reached quickly in these individuals with partial immunity, akin to treatment-associated resistance^40,41^, intensifying host-virus interactions to a degree that renders adaptive processes such as selection and recombination more readily detectable^7^.

The mutational spectrum described in this patient includes a mixture of advantageous, neutral, and deleterious changes affecting viral viability. Amplicon-based sequencing methodology identified a greater diversity of low-frequency mutations than bait capture. However, the extent to which this represented real detection of rarer genotypes, some of which may be non-viable, rather than being the consequence of sequencing noise remains unknown^42^.

Some mutations such as S98F, were detected early, whereas others likely emerged as adaptations to the host environment. V36M conferred the highest infectivity across all cell lines, with the most significant effect in low-TMPRSS2/high-ACE2 expressing cells – consistent with the Omicron lineages that rely less on TMPRSS2 and preferentially use the endocytic entry pathway^43,44^ – yet V36M was not positively selected over V36I or V36. It remained at low frequency throughout infection, illustrating that gain in fitness does not necessarily predict dominance in real-world settings likely due to complex interplay of selective pressures.

The addition of E340D enhanced infectivity across all mutations, indicative of a potential positive epistatic effect with respect to fitness, with the greatest enhancement observed in the GE.1 backbone virus. While S98F decreased infectivity in colorectal adenocarcinoma Caco-2 cells, it was also associated with higher sensitivity to serum neutralizing antibodies relative to the backbone for immunocompetent individuals. This effect was diminished by the addition of E340D, indicating the immune evasion potential of E340D in the absence of sotrovimab. A similar pattern – elevated titres relative to the backbone curtailed by the addition of E340D – was evident for V36M and V213G. To our knowledge, this represents the first documentation of a monoclonal antibody resistance mutation that simultaneously enhances both infectivity and evasion of neutralizing antibodies in absence of drug-induced selective pressure. It is important to note that while E340D does not appear to incur a fitness cost in terms of infectivity and responsive neutralizing titres, it remains rare (<0.5% cumulative prevalence worldwide as of March 2025), and E340 is highly conserved among global sequences.

SARS-CoV-2 infection downregulates ACE2 expression primarily through binding of viral S protein and the ACE2 receptor. Proposed mechanisms include clathrin-mediated endocytosis of the S-ACE2 complex leading to lysosomal degradation^45^, or endocytosis-independent pathways in which spike engagement triggers ACE2 mRNA degradation^46^. Downregulation occurs prior to viral replication, suggesting the viruses contributing to the intracellular gene pool are those that enter during the initial infection event. SARS-CoV-2 exhibits strong superinfection exclusion and reinfection of the same cell after initial infection is uncommon.

Consequently, recombination can arise in one of two ways: 1. two viruses infect the same cell before exclusion is established, or 2. a breach of the superinfection exclusion allows reinfection.

The recombination observed in the patient in this study within the ORF1ab gene reflects the exchange of genetic material between sequentially genetically distinct descendant viruses. The timing of this recombination is unknown. As such, it cannot be determined whether this recombinant virus was generated during the period that the dominant virus replacement was taking place and then persisted at low frequency until achieving dominance several weeks later, or whether the previously dominant virus persisted at low frequency and the original recombinant was generated close to the point at which its descendants became the majority viral type. Duerr et al. identified a recombinant Delta AY.45/Omicron BA.1 virus within an immunocompromised individual treated with sotrovimab post-infection during a second documented infection episode^47^. The first and second episodes occurred only two months apart and were not confirmed to be genetically distinct by sequencing. Therefore, the providence of the recombinant that they detected is uncertain. It may be that the first episode persisted subclinically, allowing recombination to occur after infection with the second variant, or that the second infection was initiated by a recombinant virion generated somewhere else. But the cases presented here and by Duerr et al, both illustrate the risks for amplification of novel recombinant viral types in immunocompromised patients, adding to the evidence that control of infection in the most vulnerable may have society-wide benefits, as well as benefits to those individuals.

In this patient, monoclonal antibodies provided a surrogate defence, as evidenced by detectable neutralizing titres across multiple timepoints prior to infection. However, the increase and subsequent fixation of E340D completely abolished this intervention, eliminating any neutralizing activity, and likely modulating the evolutionary trajectory over the course of infection. Interestingly, despite a chronic infection and the presence of E340D, the patient did not develop severe disease requiring hospitalization, a key objective of sotrovimab.

Our study has several limitations, most notably the lack of examination of non-S genes, that may exert negative or positive epistasis on the mutations identified in the S gene. Future studies should investigate mutations in genes mediating viral-host interactions as SARS-CoV-2 is highly conserved in non-structural genes encoding intra-viral protein interactions ^48^– a form of plasticity that preserves fitness while avoiding a nonviability threshold. Notably, the apparent absence of a fitness cost for E340D *in vitro* combined with it not being observed in real-world settings suggests that there may be inter-gene interaction or environmental factors required for transmission, factors that are difficult to assess in laboratory settings. In addition, at the time of the final sample collection, the patient remained SARS-CoV-2 positive, but further follow-up was not possible as the patient had completed the protocolised 48 weeks of follow-up in the trial.

A primary concern of PrEP is the potential limitation of use in those who seroconvert with a viral population harbouring low-frequency resistance mutations. To overcome the low genetic barrier to resistance for such antibodies, dual therapies can be employed; while resistance may still emerge, it generally requires more mutations than monoclonal therapy alone^49^. Alternatively, given the low prevalence of immunocompromised individuals, sequencing could guide treatment and avoid sub-optimal interventions post-hoc, although this approach is not applicable in context of PrEP. Ultimately, management of SARS-CoV-2 in immunodeficient individuals is critical to mitigate severe disease, hospitalization, and limit viral adaptation and transmission, and emergence of resistance has been documented extensively in this population^39^.

Our data suggests E340D may pose a potential risk if selected through treatment or failed prophylaxis due to its positive epistasis interactions and minimal fitness costs. However, the relative rarity of E340D is striking given its apparent phenotypic advantage, suggesting the fitness benefits of certain mutations in the context of SARS-CoV-2 may be constrained by real-world selective pressures.

## Data Availability

All data generated during this study are included in this manuscript and upon publication are available upon reasonable request to the authors.

https://doi.org/10.55876/gis8.250930sz

---

**Supplementary Figure 1.**
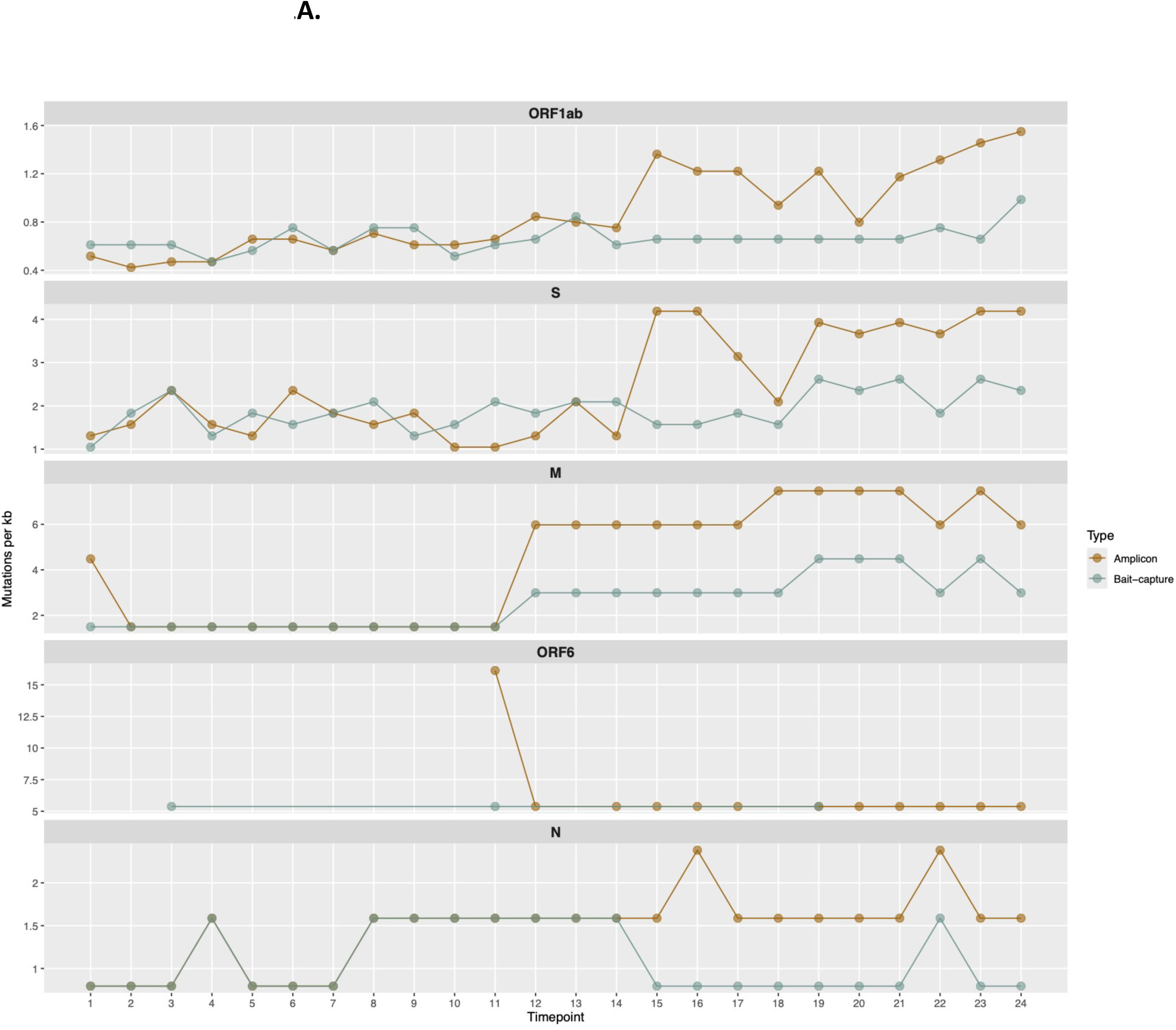
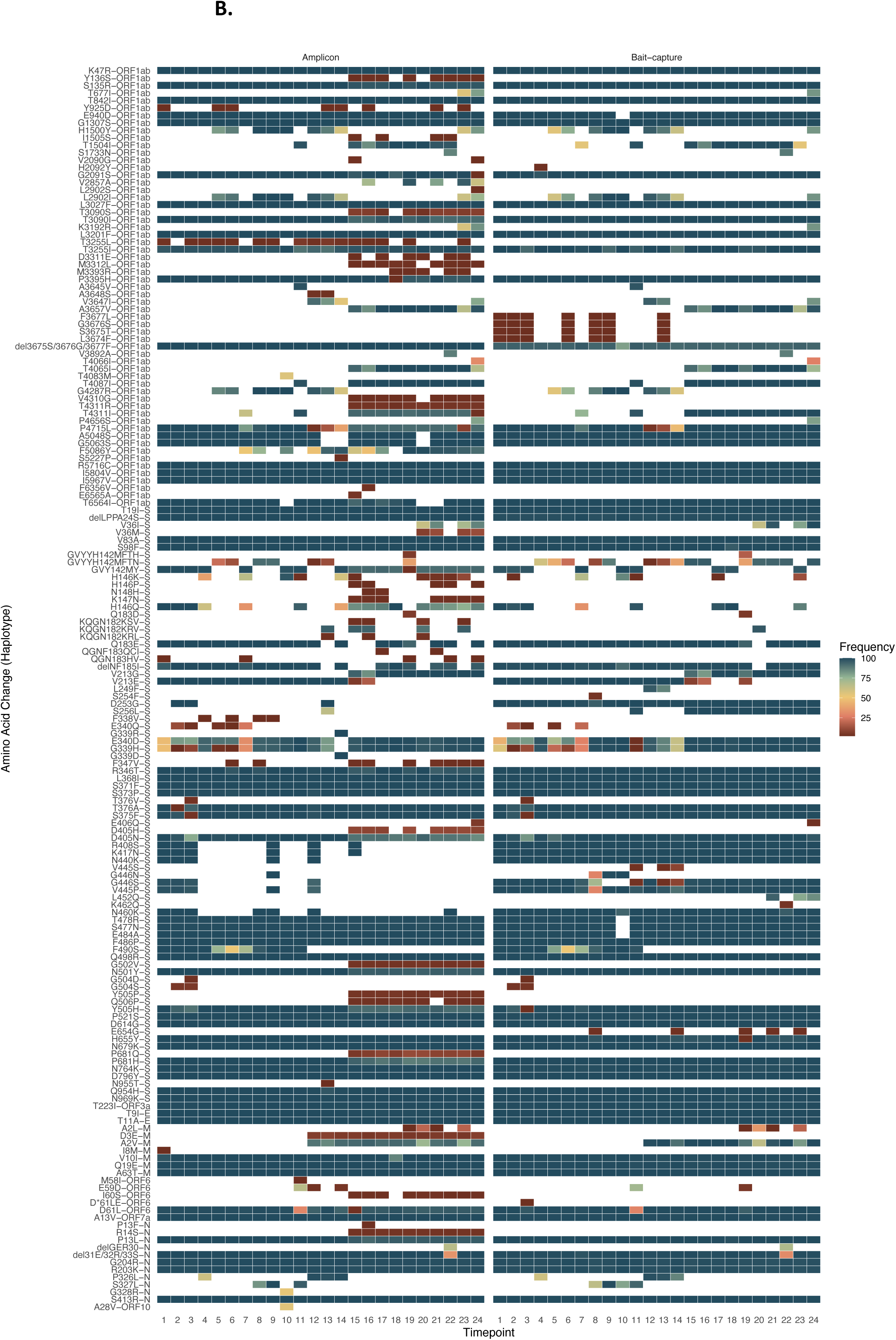
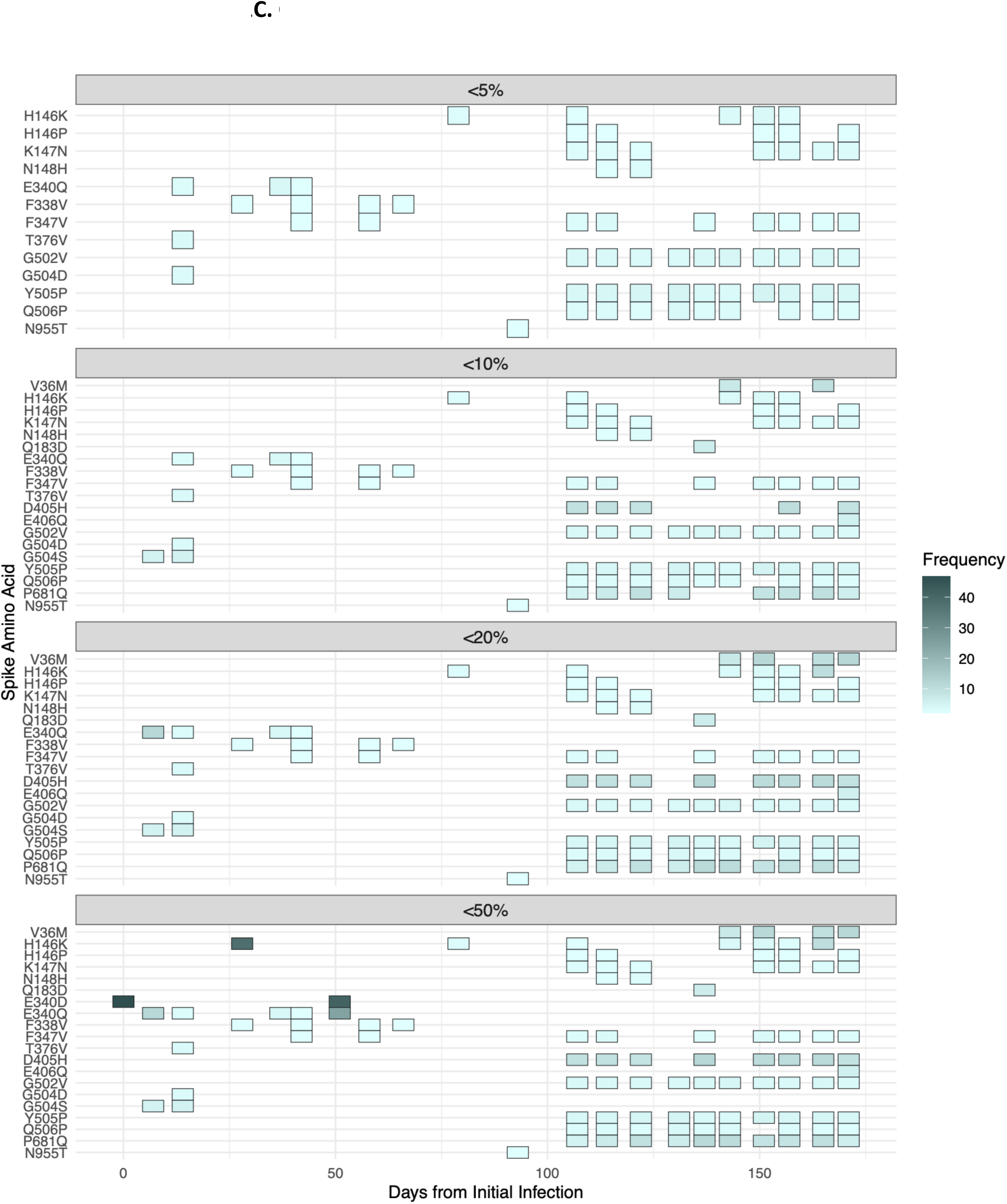
Changes in mutation density and low-frequency landscape after 100 days of infection. A) All 24 samples were sequenced through amplicon and bait-capture methods, and mutation density was assessed per kilobase. Only genes with changing temporal counts are shown. Non-lineage defining mutation counts were highest in ORF1ab (amplicon 447, bait-capture 339) and spike (S) (amplicon 228, bait-capture 175) genes. Mutation density per kilobase (kb) was highest in matrix (M) (amplicon 105, bait-capture 61), followed by ORF6 (amplicon 75, bait-capture 16), S (amplicon 60, bait-capture 46), nucleocapsid (N) (amplicon 35, bait-capture 26), and ORF1ab (amplicon 21, bait-capture 16). B) Amplicon and bait-capture sequencing exhibit high concordance. Across the full mutation spectrum, including lineage-defining mutations, both amplicon and bait-capture sequencing approaches produced largely concordant results, with bait-capture sequencing demonstrating greater genome-wide consistency. In contrast, amplicon sequencing detects a higher diversity of low-frequency (<50%) mutations. C) The low-frequency mutation landscape after 100 days of infection was characterized by unique non-consensus mutations (<50%), including mutations detected at early timepoints that persist at later timepoints despite remaining at low frequencies.

**Supplementary Figure 2.**
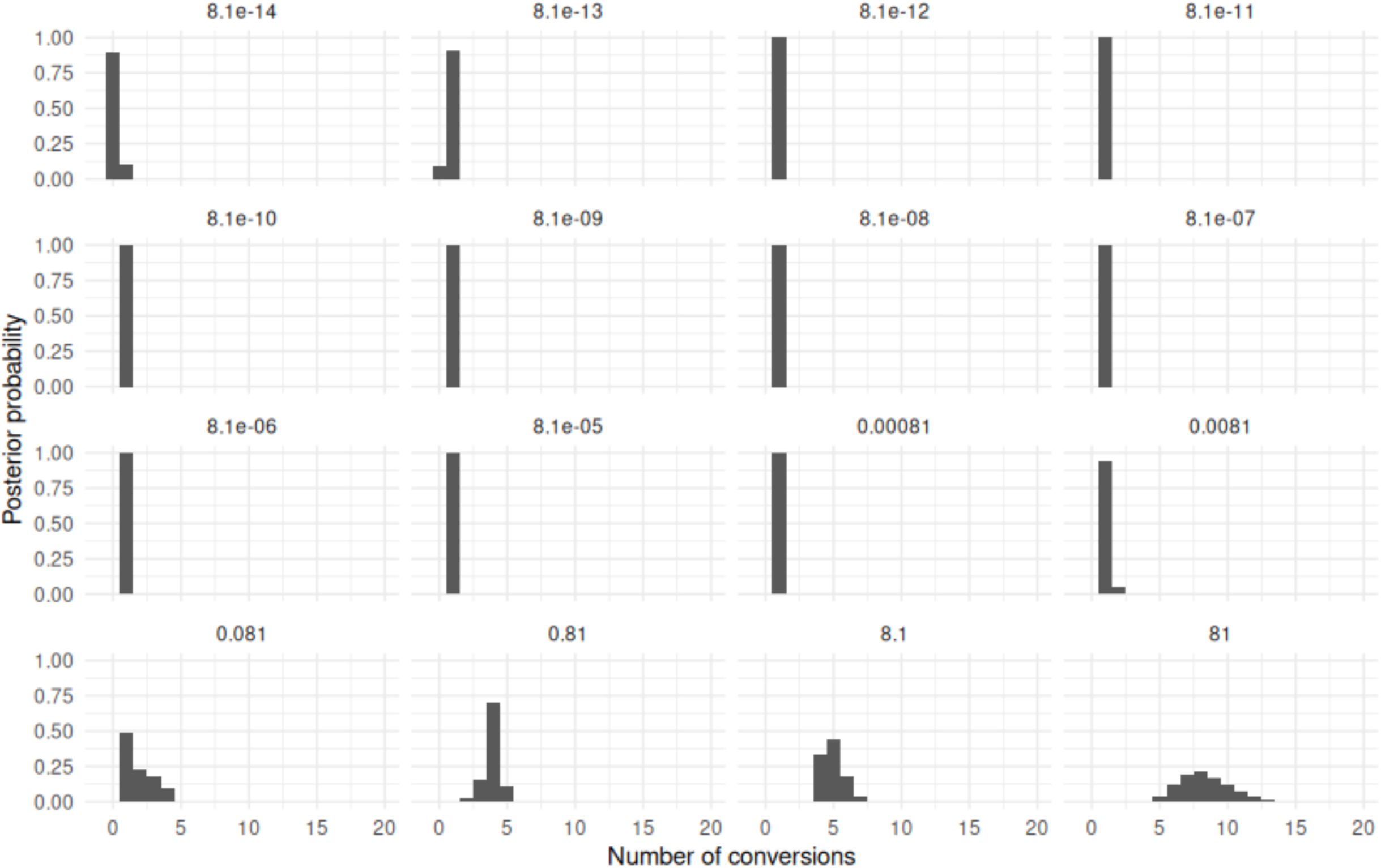
Sensitivity to the posterior number of gene conversions to the rate of gene conversion parameter. Panels are by the prior expected number of gene conversions (gene conversion rate x expected prior tree length).

**Supplementary Figure 3.**
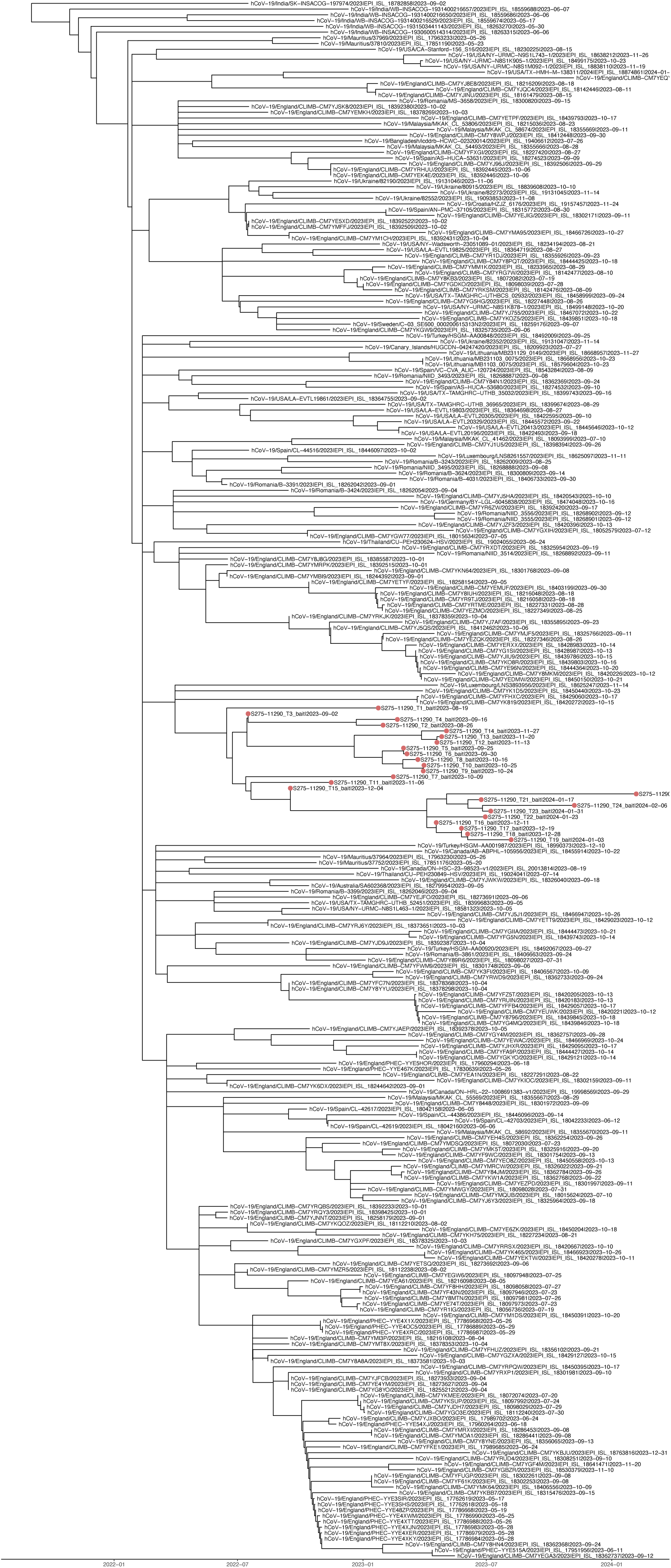
Participant sequences form distinct cluster relative to global GE.1 sequences. Maximum likelihood phylogeny inferred using IQ-TREE and temporally resolved with TreeTime display clustering of participant sequences with long branches indicative of high evolutionary divergence.

**Supplementary Figure 4.**
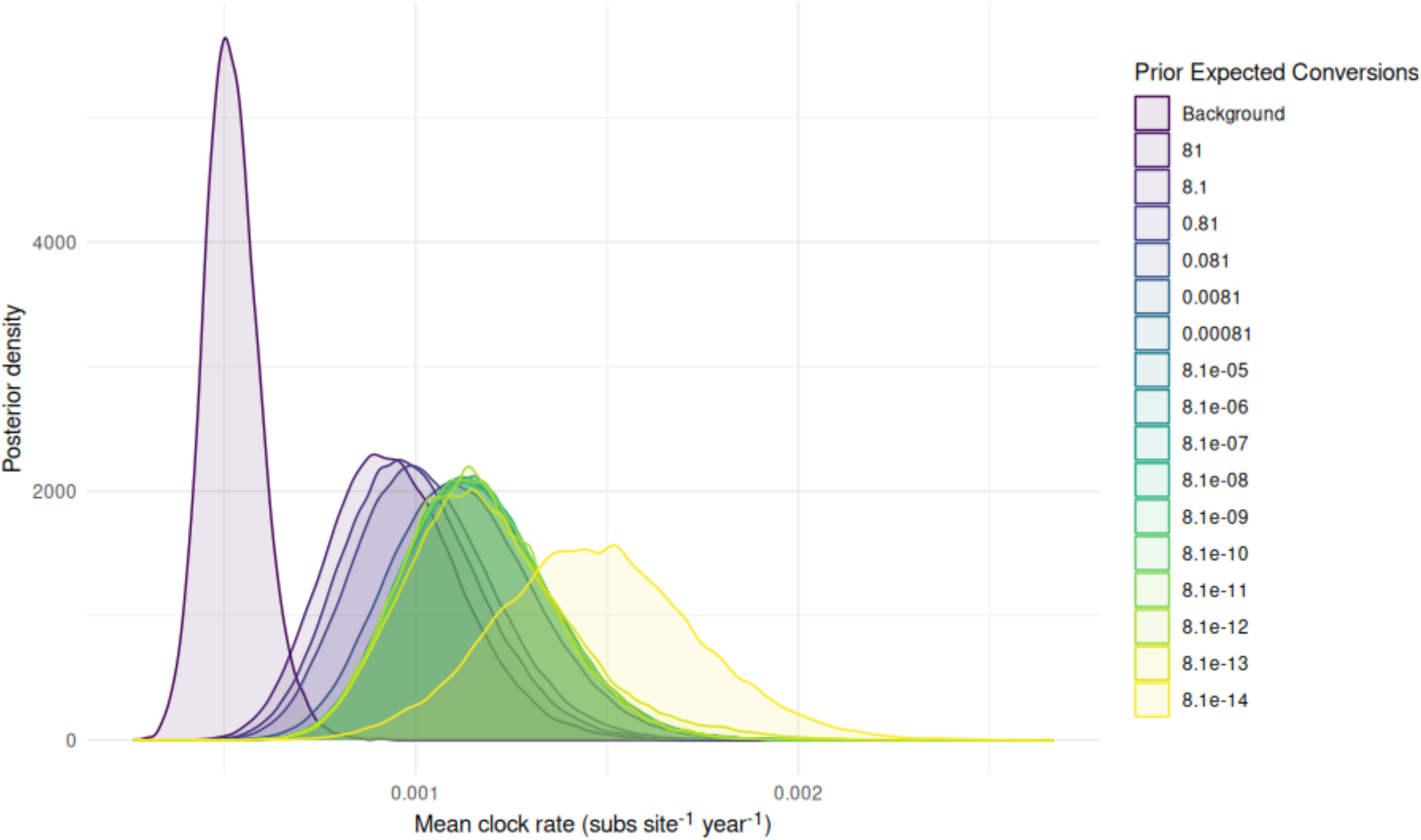
Sensitivity to the posterior mean clock rate to the rate of gene conversion parameter. Posterior distribution from the clock rate for a tree built from all GE.1 sequences on GISAID and the first sample from the patient (Background) under the GTR+R4 model with a lognormal relaxed clock and skyline tree prior and a series of trees built from the patient (Patient) under the GTR+R4 model with a strict clock and bacter tree prior and a log-linear sequence of gene conversion rates, moving the expected number of prior conversions from 81 to 8.1e-14.

**Supplementary Figure 5.**
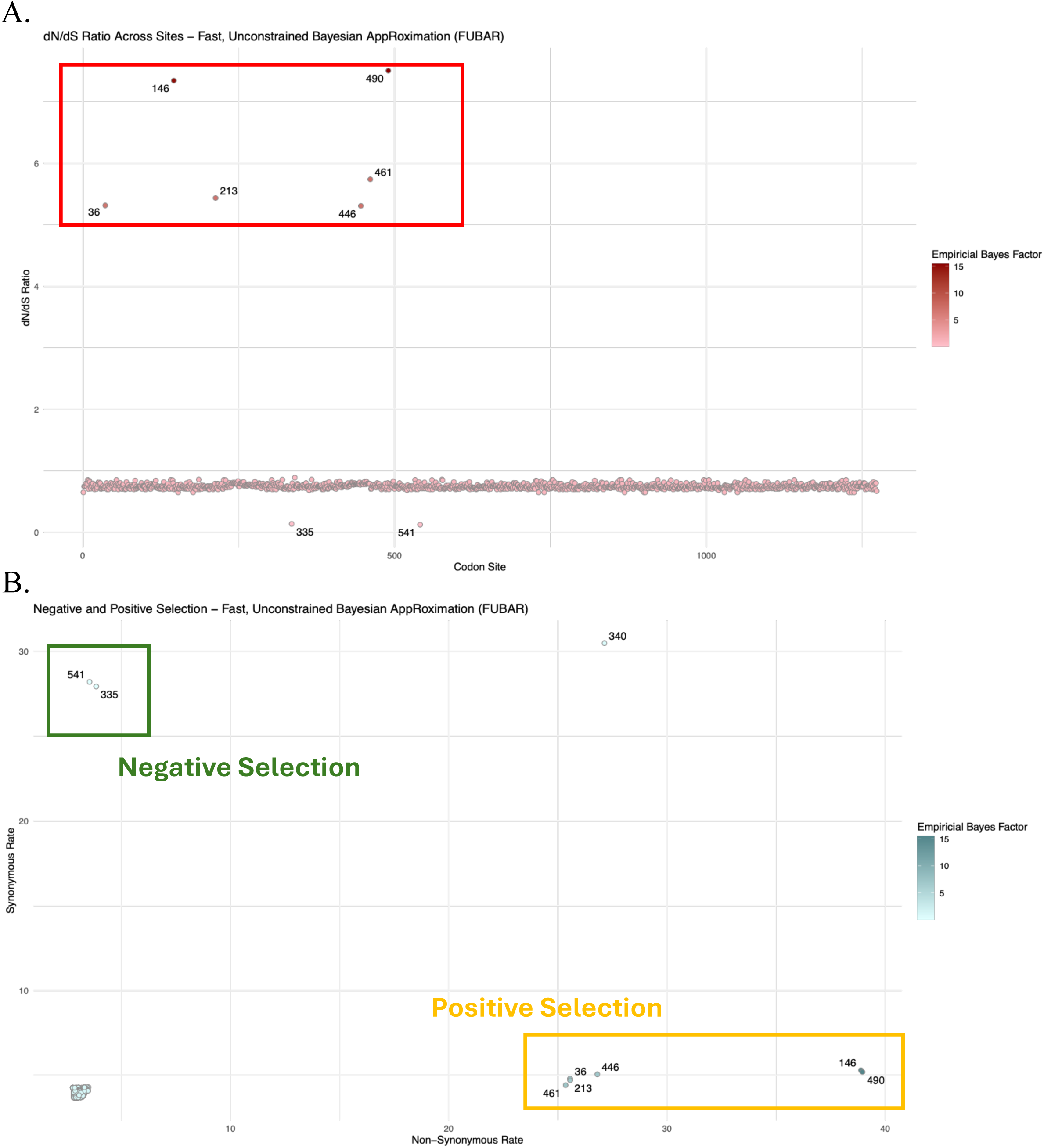
Site-specific selection in the spike gene was analyzed using FUBAR. A) The ratio of nonsynonymous (dN) to synonymous (dS) substitution rates (ω) was used to detect potential site-specific selection. Sites 36, 146, 213, 446, 461, and 490 exhibited elevated ω values, consistent with strong positive selection. B) Comparison of dN and dS substitution rates identified sites under negative selection, and a hypervariable site, E340.

**Supplementary Figure 6.**
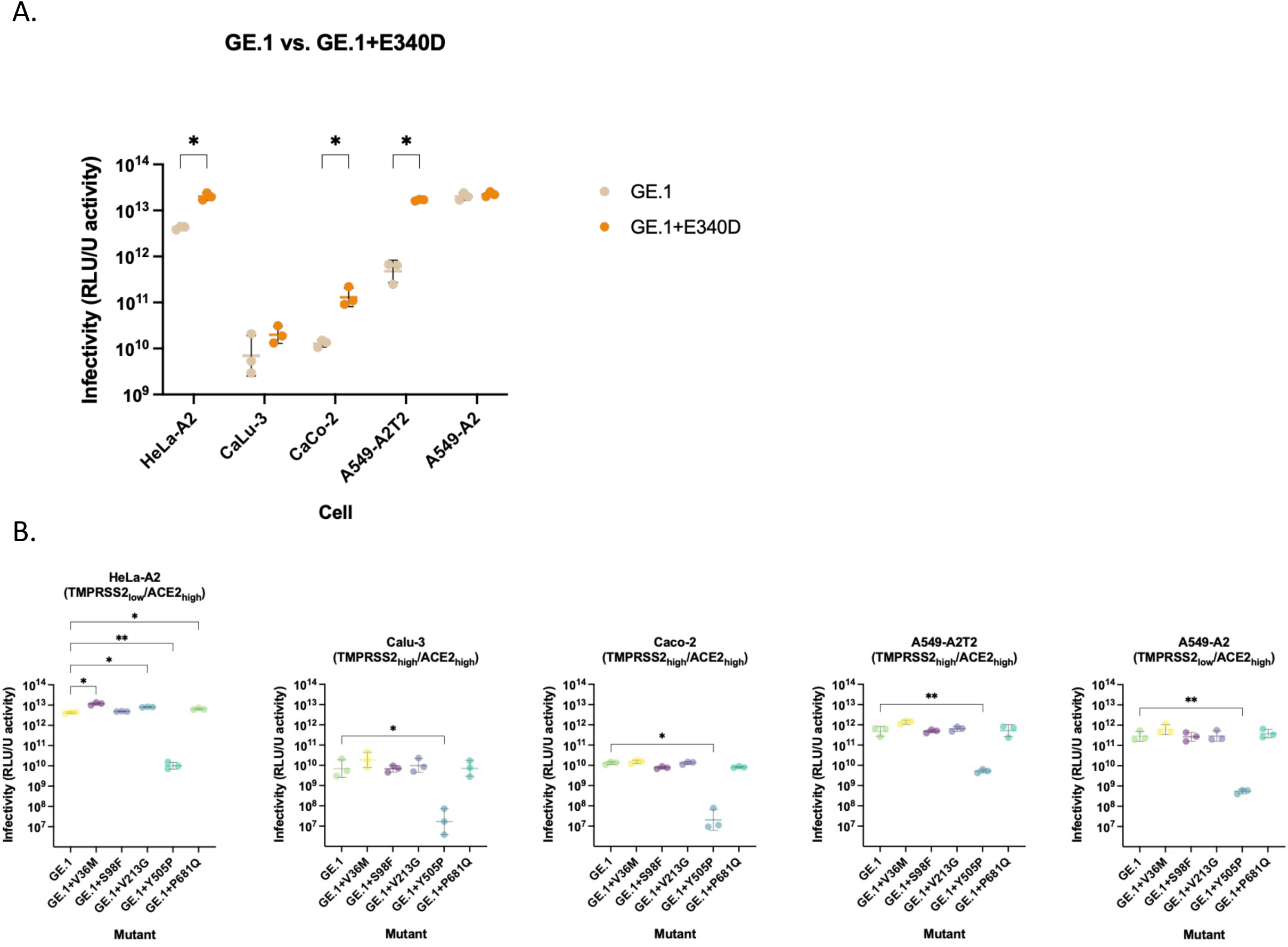
Temporal mutations positively and negatively impacted infectivity. A) The addition of E340D to the GE.1 backbone was associated with increased infectivity in HeLa-A2, Caco-2, and A549-A2T2 cells (Lognormal Welch’s t test with Bonferroni-Dunn adjustment; p-value=0.005, 0.034, and 0.039, respectively). B) Mutation Y505P decreased infectivity in HeLa-A2, Calu-3, Caco-2, A549-A2T2, A549-A2 cells and as a result was omitted from downstream functional and statistical assessment.

**Supplementary Table 1a.**
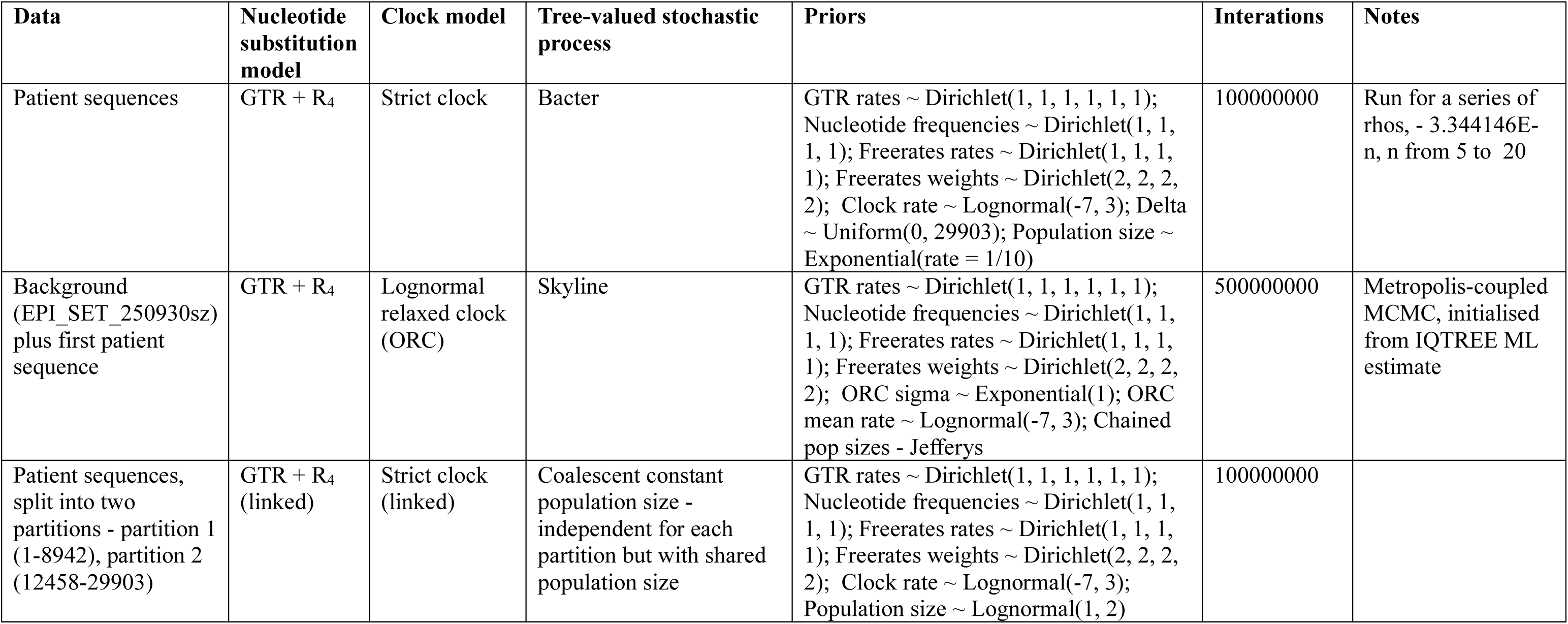

**Supplementary Table 1b.**

|  |  |
| --- | --- |
| **Data Availability** | **GISAID Identifier:** EPI_SET_250930sz<br>**DOI:** https://doi.org/10.55876/gis8.250930sz<br>All genome sequences and associated metadata in this dataset are published in GISAID’s EpiCoV database. To view the contributors of each individual sequence with details such as accession number, Virus name, Collection date, Originating Lab and Submitting Lab and the list of Authors, visit EPI_SET_250930sz |
| **Data Snapshot** | EPI_SET_250930sz is composed of 248 individual genome sequences.<br>The collection dates range from 2023-05-17 to 2024-01-22;<br>Data were collected in 19 countries and territories. |

**Supplementary Table 2.**

| **Gene** | **Type** | **Count** | **Gene length** | **Mutations per kb** |
| --- | --- | --- | --- | --- |
| M | Amplicon | 70 | 669 | 104.63 |
| M | Bait-capture | 41 | 669 | 61.29 |
| N | Amplicon | 44 | 1260 | 34.92 |
| N | Bait-capture | 33 | 1260 | 26.19 |
| ORF10 | Amplicon | 1 | 117 | 8.55 |
| ORF1ab | Amplicon | 447 | 21290 | 21 |
| ORF1ab | Bait-capture | 339 | 21290 | 15.92 |
| ORF6 | Amplicon | 14 | 186 | 75.27 |
| ORF6 | Bait-capture | 3 | 186 | 16.13 |
| S | Amplicon | 228 | 3822 | 59.65 |
| S | Bait-capture | 175 | 3822 | 45.79 |

**Supplementary Table 3.**
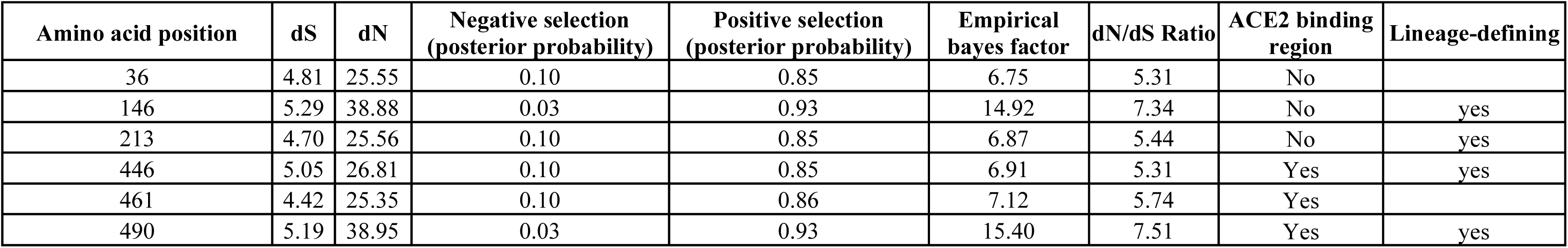

| **Amino acid position** | **dS** | **dN** | **Negative selection**<br>**(posterior probability)** | **Positive selection**<br>**(posterior probability)** | **Empirical**<br>**bayes factor** | **dN/dS Ratio** | **ACE2 binding**<br>**region** | **Lineage-defining** |
| --- | --- | --- | --- | --- | --- | --- | --- | --- |
| 36 | 4.81 | 25.55 | 0.10 | 0.85 | 6.75 | 5.31 | No |  |
| 146 | 5.29 | 38.88 | 0.03 | 0.93 | 14.92 | 7.34 | No | yes |
| 213 | 4.70 | 25.56 | 0.10 | 0.85 | 6.87 | 5.44 | No | yes |
| 446 | 5.05 | 26.81 | 0.10 | 0.85 | 6.91 | 5.31 | Yes | yes |
| 461 | 4.42 | 25.35 | 0.10 | 0.86 | 7.12 | 5.74 | Yes |  |
| 490 | 5.19 | 38.95 | 0.03 | 0.93 | 15.40 | 7.51 | Yes | yes |

**Supplementary Table 4.**

| **Variant** | **Cell line** | **Mean_log2_fc** | **Mean_infectivity_original** |
| --- | --- | --- | --- |
| GE1 | A549-A2 | 6.3 | 3.20E+11 |
| GE1 | A549-A2T2 | 5.1 | 5.20E+11 |
| GE1 | CaCo-2 | 3.3 | 1.30E+10 |
| GE1 | CaLu-3 | 1.5 | 9.74E+09 |
| GE1 | HeLa-A2 | 2.3 | 4.21E+12 |
| GE1+P681Q | A549-A2 | 5.0 | 4.17E+11 |
| GE1+P681Q | A549-A2T2 | 4.3 | 5.85E+11 |
| GE1+P681Q | CaCo-2 | 3.4 | 8.20E+09 |
| GE1+P681Q | CaLu-3 | 0.5 | 9.09E+09 |
| GE1+P681Q | HeLa-A2 | 1.4 | 6.42E+12 |
| GE1+S98F | A549-A2 | 5.3 | 2.92E+11 |
| GE1+S98F | A549-A2T2 | 4.2 | 4.90E+11 |
| GE1+S98F | CaCo-2 | 3.3 | 7.74E+09 |
| GE1+S98F | CaLu-3 | 1.1 | 7.07E+09 |
| GE1+S98F | HeLa-A2 | 1.5 | 4.88E+12 |
| GE1+V213G | A549-A2 | 4.9 | 3.29E+11 |
| GE1+V213G | A549-A2T2 | 3.2 | 6.45E+11 |
| GE1+V213G | CaCo-2 | 2.4 | 1.33E+10 |
| GE1+V213G | CaLu-3 | -0.4 | 1.21E+10 |
| GE1+V213G | HeLa-A2 | 0.5 | 7.99E+12 |
| GE1+V36M | A549-A2 | 4.6 | 6.84E+11 |
| GE1+V36M | A549-A2T2 | 3.2 | 1.38E+12 |
| GE1+V36M | CaCo-2 | 3.0 | 1.50E+10 |
| GE1+V36M | CaLu-3 | 1.1 | 2.36E+10 |
| GE1+V36M | HeLa-A2 | 0.9 | 1.17E+13 |

**Supplementary Table 5.**

| **Patient ID** | **Gender** | **Vaccine type** | **Vaccine count** | **Days post-vaccine** |
| --- | --- | --- | --- | --- |
| 1 | Male | Pfizer | 5 | 31 |
| 2 | Female | Pfizer | 6 | 30 |
| 3 | Male | Pfizer | 5 | 30 |
| 4 | Male | Moderna | 7 | 26 |
| 5 | Female | Moderna | 8 | 35 |
| 6 | Male | Moderna | 6 | 29 |
| 7 | Female | Moderna | 8 | 35 |
| 8 | Female | Moderna | 7 | 33 |

